# Single-cell spatial analysis of tumor immune architecture in diffuse large B cell lymphoma

**DOI:** 10.1101/2021.02.01.21250775

**Authors:** Anthony Colombo, Monirath Hav, Erik Gerdtsson, Mohan Singh, Alexander Xu, Alicia Gamboa, Denaly Chen, Jane Houldsworth, Rita Shaknovich, Tomohiro Aoki, Lauren Chong, Katsuyoshi Takata, Elizabeth A Chavez, Christian Steidl, James Hicks, Peter Kuhn, Imran Siddiqi, Akil Merchant

**Author notes:** **Corresponding author: Akil Merchant, MD**, Samuel Oschin Comprehensive Cancer Institute, Cedars-Sinai Medical Center 8700 Beverly Boulevard, A8103, Los Angeles, CA 90048, United States of America. Equal contribution (Anthony Colombo, Monirath Hav). **Declaration of Interests** No relevant financial interests to declare.

## Abstract

Multiplexed immune cell profiling of the tumor microenvironment (TME) in cancer has improved our understanding of cancer immunology, but complex spatial analyses of tumor-immune interactions in lymphoma are lacking. Here we used imaging mass cytometry (IMC) on 33 cases of diffuse large B cell lymphoma (DLBCL) to characterize tumor and immune cell architecture and correlate it to clinicopathological features such as cell of origin, gene mutations, and responsiveness to chemotherapy. To understand the poor response of DLBCL to immune check point inhibitors (ICI), we compared our results to IMC data from Hodgkin lymphoma (HL), a cancer highly responsive to ICI, and observed differences in the expression of PD-L1, PD-1, and TIM-3. We created a spatial classification of tumor cells and identified sub-regions of immune activation, immune suppression, and immune exclusion within the topology of DLBCL. Finally, the spatial analysis allowed us to identify markers such as CXCR3, which are associated with penetration of immune cells into immune desert regions, with important implications for engineered cellular therapies.

**SIGNIFICANCE:** This is the first study to integrate tumor mutational profiling, cell of origin classification, and multiplexed immuno-phenotyping of the TME into a spatial analysis of DLBCL at the single cell level. We demonstrate that, far from being histo-pathologically monotonous, DLBCL has a complex tumor architecture, and that changes in tumor topology can be correlated with clinically relevant features. This analysis identifies candidate biomarkers and therapeutic targets such as TIM-3, CCR4, and CXCR3 that are relevant for combination treatment strategies in immuno-oncology and cellular therapies such as CAR-T cells.

## INTRODUCTION

Diffuse large B-cell lymphoma (DLBCL) is the most common subtype of non-Hodgkin lymphoma. Although many patients are cured with standard chemo-immunotherapy, up to 40% of DLBCL patients have refractory disease or develop relapse following R-CHOP, or similar regimens, warranting the development of novel, more effective therapeutic strategies for these patients (1). The composition of the tumor microenvironment (TME) has emerged as an important predictor of DLBCL outcome in gene expression profiling studies (2–5). Pileri et al. and a few other studies have reported that higher proportion of CD4 T cells, dendritic cells, and myofibroblasts predicted better outcome in DLBCL treated with R-CHOP (4,6). Other studies reported that increased infiltration of CD8 T cells was associated with better outcome in DLBCL(7). The role of immune cells such as macrophages and regulatory T cells (T_REG_) is less clear, with contradictory reports in the literature (8–11). Despite recent methodological advances to study the TME in cancer, highly multiplexed immunophenotypic characterization of the tumor immune architecture in DLBCL has not been reported (12–15).

The two major molecular sub-types of DLBCL based on cell of origin (COO) are germinal center B-cell (GCB), originating from B-cells in the light-zone, and activated B-cell (ABC), derived from B-cells which have migrated out of the germinal center and committed to differentiation (16). Generally, GCB sub-type tends to have better overall survival compared to ABC (17,18). The COO classifications were originally performed using gene expression and have found widespread clinical application through immunohistochemistry using the HANS algorithm, which identifies GCB and non-GCB subtypes. Other aggressive sub-types include double-expressor, which overexpress *MYC* and *BCL2*, and double-hit lymphoma, which have chromosomal rearrangements of *MYC* and *BCL2*, or seldomly *BCL6* (19). DLBCL have been further subset using numerous somatic copy number alterations, structural variants and mutations which stratified DLBCL into four or five genetic subtypes (20,21). Chapuy et al. identified five distinct genetic subtypes involving top ranking mutations such as *BCL6* (coordinate genetic signature 1, or C1), *TP53* (C2), *BCL2* (C3), *SGK1* (C4), and *CD79B* (C5). Integration of mutational analysis with functional and spatial parameters derived from DLBCL TME analyses, however, has not yet been reported.

Programmed cell death receptor 1 ligand (PD-L1) is a member of the B7 family that is expressed on tumor cells and has been reported as a predictor of poor survival in multiple epithelial and hematologic malignancies including DLBCL(22–25). Blockade of PD-1/PD-L1 signaling with monoclonal antibodies such as nivolumab and pembrolizumab in patients with relapsed/refractory Hodgkin lymphoma has resulted in high and durable clinical response rates (26–28). Unlike in Hodgkin lymphoma, the response rate to PD-1/PD-L1 blockade in relapsed/refractory DLBCL (R/R DLBCL) has been disappointingly low (≤10%), even though PD-L1 is expressed in a significant subset of DLBCL and is associated with poor prognosis(29). The reasons for this discrepancy between PD-L1 expression and the failure of PD-1/PD-L1 pathway inhibition in R/R DLBCL represent a major knowledge gap in the field.

In this study, we characterize the TME of DLBCL, including the cell types, frequency, functional state and spatial topology using imaging mass cytometry (IMC)(30). Using our previous study of Hodgkin lymphoma, we identify key differences between the Hodgkin lymphoma and DLBCL TME components that might explain their differential responses to immune check point inhibitor (ICI) therapy (31). Finally, we perform spatial analysis of the DLBCL tumor architecture to reveal immunologic sub-regions of tumor immune interaction at single cell resolution.

## RESULTS

### IMC analysis of DLBCL TME identifies heterogeneity of immune infiltration and cellular subtypes

To comprehensively quantify the cellular and spatial heterogeneity in DLBCL, we designed an IMC panel to broadly cover T cells, B cells, Macrophages and structural markers (Table S1). We performed IMC on 41 representative tumor samples from 33 cases which were selected from a previously described cohort based on adequacy of residual tissue for analysis (Figure S1) (32). Our panel of 32 antibodies quantified exhaustion (TIM-3, LAG-3, PD-1) and proliferation (Ki67) states and included markers of immune and endothelial cell lineages. These findings were correlated with clinical features such ascell-of-origin (COO) by IHC HANS criteria, presence of mutations, and responses to chemotherapy (Table 1, Figure S2). In the cohort, 22 (67%) of the patients achieved a sustained complete response (CR) to rituximab based chemoimmunotherapy, while 7 (21%) were primary refractory or relapsed within one year (REF). Four patients did not receive chemotherapy and were excluded from response to therapy analysis.

**Table 1.** Clinicopathological characteristics of the cohort.

| Parameters | n (%) | Parameters | n (%) |
| --- | --- | --- | --- |
| Age, mean ± SD (range) | 49 ± 15 (20-82) | Mutation status | 22 (67%) |
| Sex |  | TP53 | 8 (24%) |
| F | 9 (27%) | BCL2 | 3 (9%) |
| M | 24 (73%) | BCL6 | 2 (6%) |
| Ethnicity |  | CD79b | 6 (18%) |
| Black | 1 (3%) | SGK1 | 1 (3%) |
| Asian | 5 (15%) | MYD88.L265P | 2 (6%) |
| Caucasian | 2 (6%) | NOTCH1 | 3 (9%) |
| Hispanic | 25 (76%) | NOTCH2 | 3 (9%) |
| LDH |  | EZH2 | 4 (12%) |
| Not elevated | 16 (49%) | 9p24 chromosomal gain | 8 (24%) |
| Elevated | 17 (51%) | Stage |  |
| IPI score |  | I/II | 15 (45%) |
| 0-2 | 23 (70%) | III/IV | 18 (55%) |
| 3-5 | 10 (30%) | III/IV | 18 (55%) |
| EBER |  | Response status |  |
| Positive | 6 (18%) | CR | 22 (67%) |
| Negative | 27 (82%) | REF | 7 (21%) |
| Hans COO |  | Unknown | 4 (12%) |
| GCB | 17 (51%) | Treatment |  |
| Non-GCB | 16 (49%) | Chemo-immunotherapy | 29 (88%) |
|  |  | Not done/LTF | 4 (12%) |

COO: cell of origin; GCB: Germinal Center B Cell; Non-GCB: Non-Germinal Center B Cell; CR: Complete Response; REF: Refractory to treatment; LTF: Lost to follow up; EBER: Epstein-Barr virus encoded small RNAs; LDH: Lactate dehydrogenase serum levels. Double-hit tumors were excluded from this data set.

IMC produces images similar to immunohistochemistry or immunofluorescence with the added advantage of increased multiplex staining. The images were segmented using pixel classification training into single cells yielding on average 16,889 cells per region of interest (ROI), as previously described (Figure S3) (33–35). Cellular expression of cell lineage, inducible states, and spatial features were hierarchically clustered with Phenograph to identify major phenotype clusters such as endothelial, regulatory (T_REG_), CD8 T cells, CD4 T cells, macrophages, and B cell tumors (Figure 1A) (36).

**Figure 1.**
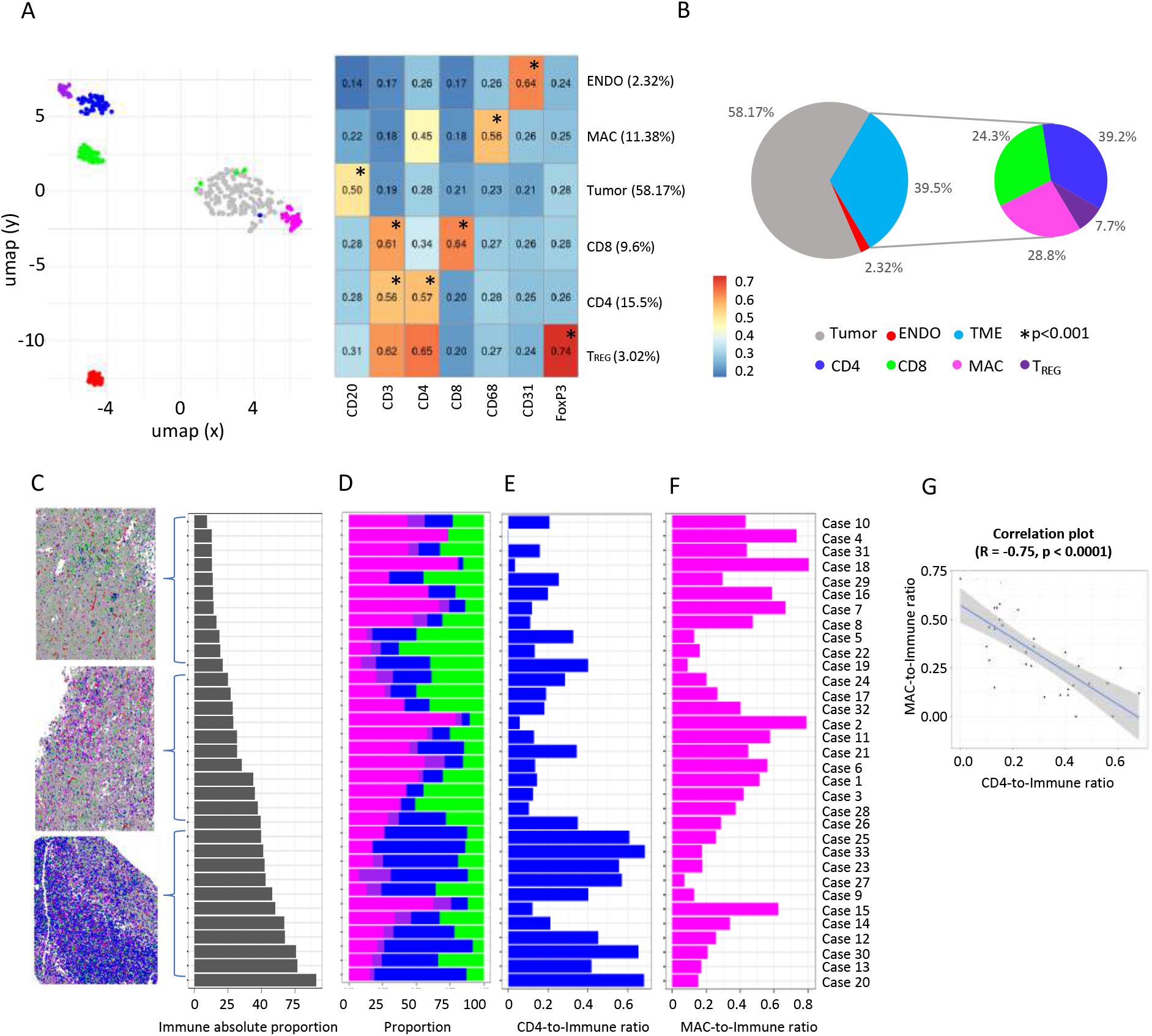
IMC analysis of DLBCL TME identifies heterogeneity of immune infiltration and cellular subtypes. A. Heatmap and UMAP of tumor, CD4, CD8, T_REG_, macrophage and endothelial meta-clusters, generated by Phenograph clustering the centroids of each subpopulation. The complete single-cell network embedding is depicted in Figure S4. Statistical testing for marker enrichment denoted with * (p<0.001, ANOVA). B. Of the 39.5% of cells comprising the TME, the composition was dominated by CD4, CD8, and macrophages, which make up 92.3% of the immune microenvironment. C. Left: Initial pathologist review revealed various degrees of immune infiltrate in DLBCL. Pseudo-colored images representative of cases with low (top), medium (middle) and high (bottom) degree of immune infiltrate. Right: Cases ranked in order of absolute proportion of immune cells (9.42% to 90.14%). See Figure S7 for all cases D. Analysis of the TME composition showed marked heterogeneity in the distribution of CD4, CD8, T_REG_ and macrophages across cases. E. The proportion of CD4 increases with the increasing proportion of immune infiltrate. F. The proportion of macrophages decreases with the increasing proportion of immune infiltrate. G. Negative correlation between the proportion of CD4 T cells and that of macrophages.

Hierarchical meta-clustering across ROIs first identified 14 meta-clusters which were well distributed across cases (Figure S4). The initial meta-clusters were constructed and annotated based on lineage marker expression which identified the broadest categories of the TME phenotypes in DLBCL(36). Quality control analysis ensured that cluster annotations were appropriately defined (Figure S5, and S6). The major cell components across the 33 cases included 58.2% malignant B-cells, 39.5% TME, and 2.32% endothelial cells (Figure 1B). Within the TME, we identified 7.70% T_REG_, 24.3% CD8 T cells, 28.8% macrophages and 39.2% CD4 T cells. Similar to previous reports, macrophage proportions were 3.96% (p=0.069) higher in non-GCB compared to GCB cases(37). In tumors with low immune infiltration, (Figures 1C, 1D, and S7) macrophages were the predominant immune cell type, whereas in cases with greater immune infiltrate CD4+ cells predominated (Figures 1E and 1F). This resulted in a significant negative association between the macrophage and CD4+ cell proportions (Pearson’s R=-0.75, p<0.001) across the 33 cases (Figure 1G) as was similarly reported for solid tumors (12).

### Association between cell of origin and abundance of cell phenotypes in DLBCL TME

We next performed sub-classification of each major cell component by including inducible markers and morphological features, which identified the functional states of tumor and immune sub-phenotypes in the TME (Figure 2A). We then associated these TME sub-clusters with IPI, and the five Chapuy genetic subtypes (C1-C5) and cell-of-origin by Hans classification to gain insight into the biological significance of the observed tumor and immune heterogeneity.

**Figure 2.**
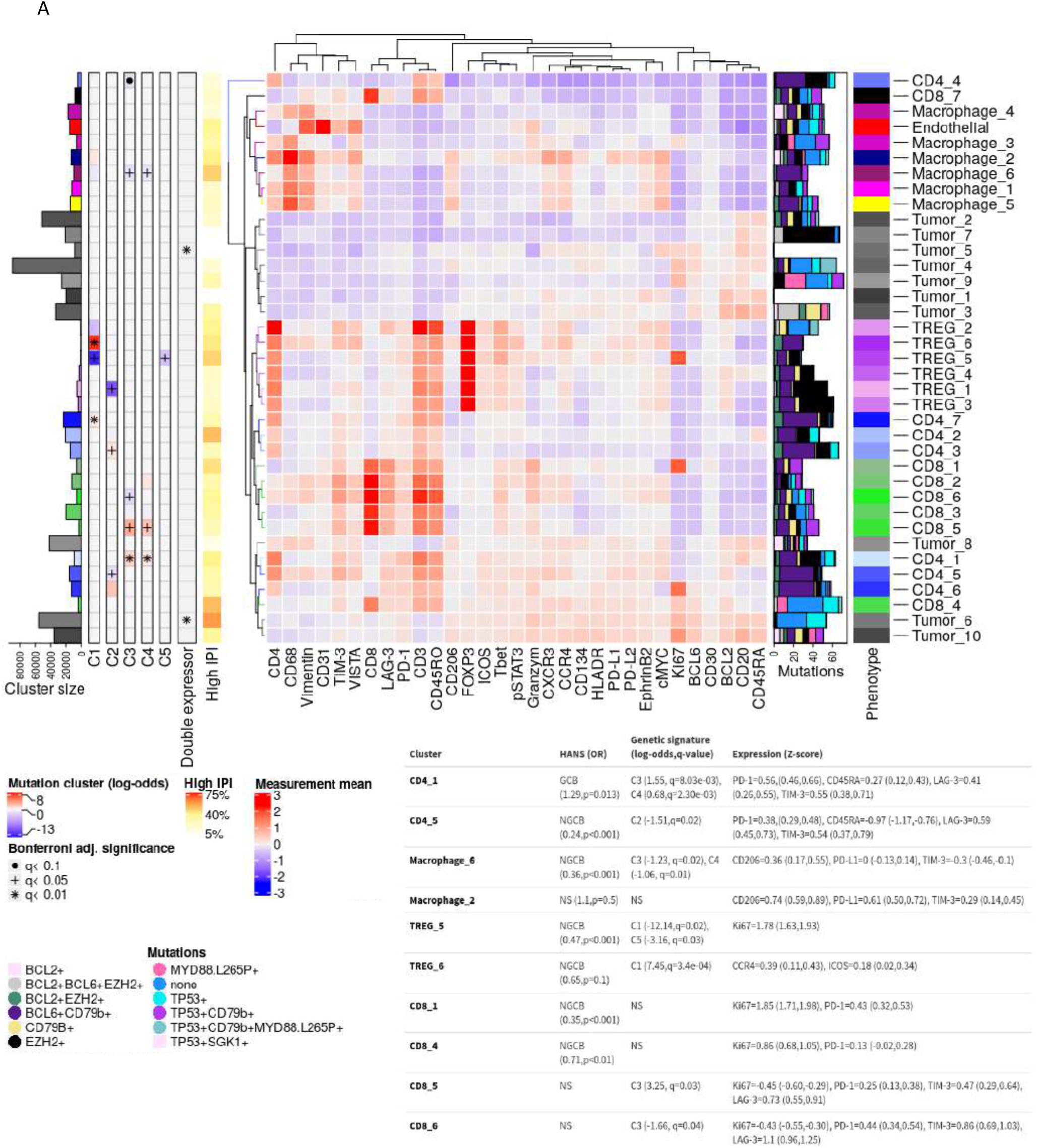
Association between genetic mutations, cell of origin and abundance of sub-cellular phenotypes in DLBCL TME. Sub-phenotypes were created by re-clustering cells using all markers. Heatmap depicts heterogeneity of clinical predictors, genetic mutation signature and protein expression across all tumor and immune sub-phenotypes. The cluster sizes, strength of association of expression clusters and the cluster proportion of patient ‘high’ IPI scores greater than 3 portrayed as a cluster percentage. The sub-phenotype log-odds of association to somatic copy number alteration genetic signature classes (C1-C5) are depicted using Bonferroni adjusted significance. Clusters significantly co-expressing cMYC/BCL2 or cMYC/BCL6 (p<0.01; mixed-effects ANOVA) were determined as ‘double expressor’ based on average normalized cellular intensity. Genetic mutations relevant to DLBCL were included as a cluster proportion (%) depicted on the right.

The CD4 sub-cluster 1, or CD4_1 was associated with genetic subtype C3 and C4. CD4_1 had a 371% increase in odds of the genetic subtype C3 for each percent increase in CD4_1 proportion (Bonferroni adj. P<0.01) and 97% odds increase in C4 (Bonf. adj. P<0.01). CD4_1 was enriched for GCB (Chi-square ORGCB=1.29, p=0.013) which was concordant with the C3/C4 coordinate genetic signatures found in primary GCB tumor subtype(20). CD4_1 had the highest PD-1 mean normalized intensity (p<0.001) (Figure S8A). CD4_1 was labeled as the CD45RA^+^ exhausted phenotype because of its high levels of CD45RA, LAG-3 (p<0.001), and TIM-3 (p<0.001)(Figure S3). In contrast, CD4_5 was negatively associated with genetic subtype C2 and each percent increase in the CD4_5 reduced the odds of C2 by 78% (Bonf. Adj. P=0.02). CD4_5 was significantly associated with non-GCB (ORGCB =0.24, p<0.001) and was defined as the CD45RA^−^ exhausted phenotype due to its low CD45RA intensity (p<0.001), and increased expression of LAG-3 (p<0.001) and TIM-3 (p<0.001). Across patients, CD4 sub-clusters had significant expression variability of CXCR3, CCR4, PD-1, PD-L1, ICOS, Ki67, and VISTA. While the limited numbers of patients in this study prevented formal testing of these markers for predictive value, their heterogeneity across samples makes them intriguing candidate biomarkers.

The M1 macrophage (Macrophage_3, Macrophage_4) phenotype family was not associated with either COO type, whereas the CD206 expressing M2 (Macrophage_1, Macrophage_5, and Macrophage_6) family was enriched with NGCB (ORGCB=0.39, p<0.001). When examining the association with genetic subtypes, Macrophage_6 was negatively associated with C3 and C4 with a 71% (Bonf. Adj. P=0.02) and 65% (Bonf. Adj. P=0.01) relative decrease odds for each percent increase in Macrophage_6, and is consistent with C3/C4 DLBCL being primarily of GCB cell-of-origin.

T_REG__2 and T_REG__6 were grouped together as “highly suppressive T_REG_” because of high expression of CCR4 (p<0.001) and ICOS (p=0.02). Highly suppressive T_REG_ were marginally represented in NGCB subtypes (ORGCB=0.76, p=0.08). However T_REG__6 was strongly associated with genetic subtype C1 and for each percent increase in T_REG__6, the odds of C1 genetic subtype increased 1719% (Bonf. Adj. P<0.001). In contrast the T_REG__5 cluster, or Ki67^+^T_REG,_ had an activated proliferative phenotype with high Ki67 (p<0.001), and was strongly associated with non-GCB (ORGCB=0.47, p<0.001). Interestingly, T_REG__5 was negatively associated with genetic subtypes C1 and C5 with a 99% (Bonf. Adj.P=0.02) and 95% reduction (Bonf. Adj.P=0.03), respectively, for each percent increase in T_REG__5, which was unexpected as genetic subtypes C1/C5 are primarily ABC subtype(20). These findings suggest that TME findings may reveal additional heterogeneity not captured by COO or genetic subtypes.

The CD8 sub-clusters had significant variability in activation and exhaustion markers such as TIM-3, PD-1, and LAG-3. CD8_1 and CD8_4 had an Ki67+ activated phenotype (p<0.001) and high PD-1 expression ((p<0.001), and were strongly associated with non-GCB (ORGCB =0.48, p<0.001) however, the Ki67^+^CD8 phenotype did not have significant associations with the DLBCL genetic subtypes. Although this population was PD-1^+^, it had low expression of other exhaustion markers consistent with an activated phenotype. In contrast several CD8 clusters demonstrated an exhausted non-proliferative phenotype with co-expression of PD-1, LAG-3 and TIM-3. For example CD8_6 showed a negative association with genetic subtype C3 with a 81% reduction (Bonf. Adj. P=0.04), while CD8_5 showed a strong positive association with C3 with an increased 248% (Bonf. Adj. P=0.03) for each percent change of the CD8 cluster. While these clusters show similar expression of exhaustion markers, CD8_6 had higher expression of chemokine receptors CXCR3 and CCR4 compared to CD_5 suggesting differences in chemokine gradients and immune trafficking. CD8_2 also shared a similar exhausted phenotype however it showed a strong enrichment in NGCB tumors (ORGCB =0.42, p<0.001). Interestingly this CD8 population had higher pSTAT3 activity comparted to other exhausted sub-types. These results demonstrate marked heterogeneity in the CD8 populations revealed by IMC. Importantly, we show that both activated and exhausted populations expressed PD-1, demonstrating the limitations in using PD-1 as a single marker for exhausted T cells in tissue sections.

### Association between chromosomal abnormalities and PD-L1 expression in DLBCL TME

Each patient’s chromosomal abnormalities were evaluated using comparative genomic hybridization (CGH), and we observed that 9p24 copy gains or losses were not associated with tumor abundances (p=0.58, ANOVA) and marginally associated with tumor PD-L1 expression (p=0.21, ANOVA), although this is likely due to the limited sample size as 9p24 copy gains have been previously associated with PD-L1 expression in this cohort (32). Although our IMC panel was primary focused on the TME and did not include the markers needed for COO classification by HANS, the immuno-morphological tumor clusters identified by IMC were aligned well with mutational clusters and COO classification. Follow up work using a tumor focused panel of markers would likely identify additional tumor heterogeneity.

### Association between chemo-responsiveness, tumor phenotypes, and TME

In order to understand how tumor and TME interactions contribute to chemo-responsiveness, we examined the abundance and spatial interactions of tumor and immune cell clusters in relation to clinical response to chemotherapy. To examine changes in the TME we computed a generalized estimating equation model, and for a percent increase in M1-Macrophage case proportion there was a 79.2% reduction in the odds of treatment refractory (log-odds=-1.57, Bonferroni adjusted p-value=0.02)(Figure 3A).

**Figure 3.**
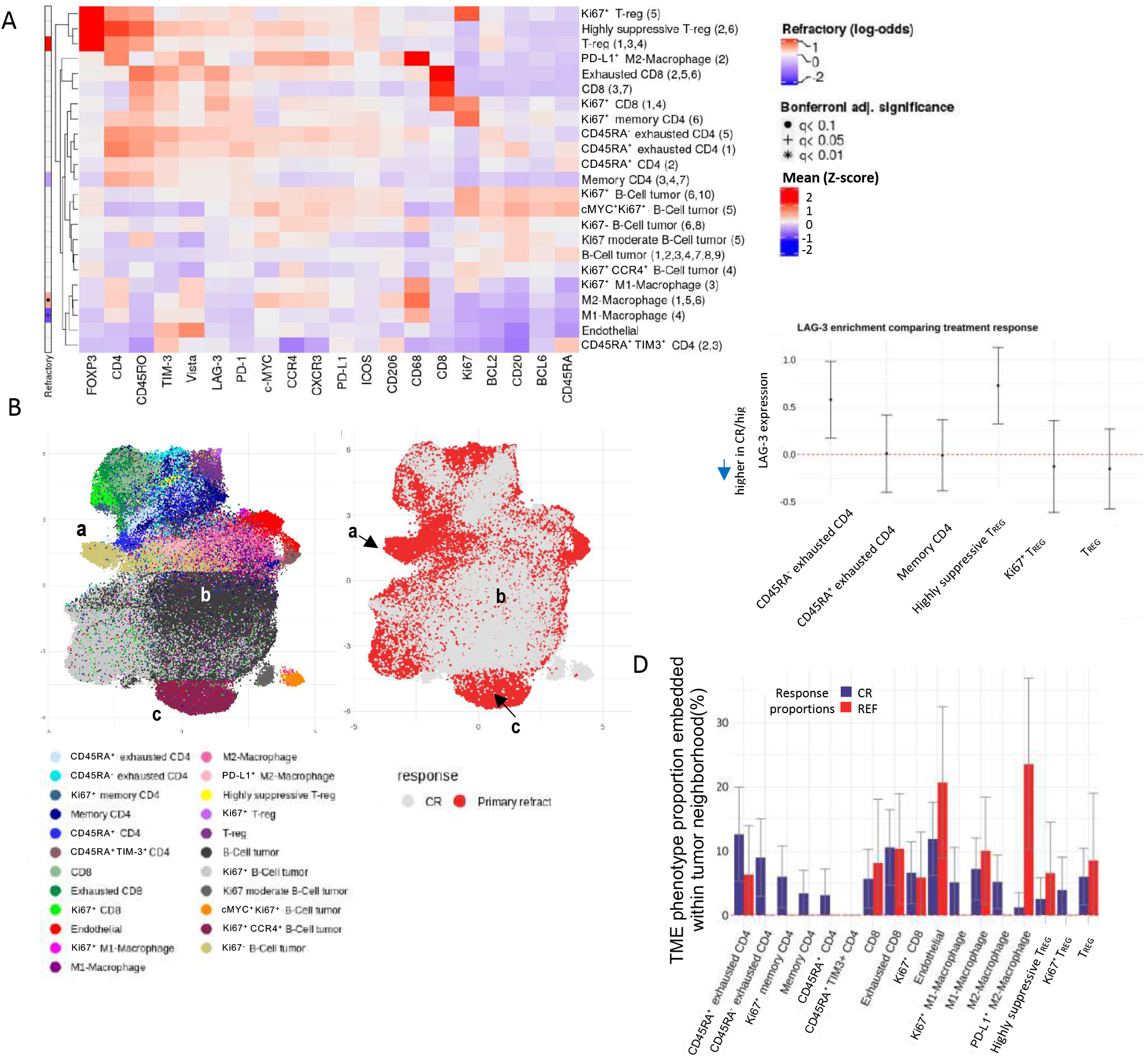
Spatial arrangement and enrichment of tumor subsets in the context of chemotherapy response status. A. The annotated phenotype expression profile is depicted using the mean normalized intensity (z-score). (Left) Annotation plot depicts the multivariate generalized linear model which identified M1-macrophages with significantly decreased log-odds for refractory (Bonferroni adjusted p-value<0.05), whereas M2-macrophages had marginally increased log-odds for refractory (Bonferroni adjusted p-value<0.09). B. The (left) UMAP projects the structure of each cluster, and adjacent (right) UMAP depicts the enrichment of phenotypes in treatment response. The enriched refractory tumor phenotypes are Ki67-(a), and Ki67+CCR4+ B cell tumor (c), whereas the B cell tumor phenotype enriched in complete responders are denoted as (b). See Figure S9A. C. The global ANOVA model comparing LAG-3 expression (z-score) on each immune subset comparing refractory (REF) subjects to complete responders (CR). The average intensity differences are depicted on the y-axis with 95% confidence interval. The CD45RA^−^ exhausted CD4 LAG-3 average intensity was higher in REF compared to CR (p=0.10), and similarly in highly suppressive T_REG_ (p=9.2e-03). Above the dashed line indicates higher normalized expression in refractory subjects, and below indicates increase in responders. See Figure S9B D. In order to understand chemo-refractoriness at the hyper-local TME level, spatial interactions of immune phenotypes within 2-3 cell diameters (15 µm) of the tumor cells were computed using 1,000 permutations (p<0.025). The relative proportions (%) of significant spatial interactions of immune phenotypes within the tumor neighborhoods are depicted on the y-axis. Within refractory tumor neighborhoods, PD-L1^+^M2 macrophages had increased co-localization compared to tumor neighborhoods in complete responders and CD4 subsets were co-localized with CR.

We then performed Uniform Manifold Approximation and Projection (UMAP) of all samples to visualize distinct clusters of tumor and immune cells (Figure 3B, S9A). In particular, we were interested in understanding heterogeneity of PD-L1 expression on tumor cells as this has been previous associated with poor prognosis after chemotherapy(22,23,25). The largest cluster (67.9%) of tumor cell was labeled “B cell tumor” (region b) and was enriched in complete responders and did not express PD-L1. Tumor regions c and a were enriched in refractory patients. Cluster a was identified as Ki67-tumor and had high PD-L1 expression (PD-L1 estimate=+0.33, 95% CI: (0.22,0.43), p<0.001) while cluster c was identified as Ki67^+^CCR4^+^ tumor and had low PD-L1 expression (PD-L1 estimate=-0.21, 95% CI: (−0.32, −0.09), p<0.001) These data confirm that PD-L1 expression was associated with refractoriness to chemotherapy, but that its expression was limited to a certain sub-population of non-proliferating tumor cells. This finding suggests the interesting possibility that chemoresistance is mediated by a non-proliferating, immune evading subpopulation of tumor cells.

We then examined the TME for markers that associated with chemotherapy response. Comparing refractory to complete responder average intensity, we observed that LAG-3 was increased in REF subjects, specifically in the CD45RA^−^ CD4 exhausted phenotype (LAG-3 difference estimate=0.58, 95% CI: (0.18, 0.98), p=0.10) and significantly increased in highly suppressive T_REG_ (LAG-3 difference estimate=0.72, 95% CI: (0.32, 1.13),p=9.1e-03) (Figure 3C). Additionally, both phenotype case proportions did not differ between REF to CR, although LAG-3 expression was enriched in REF subjects (Figure 3C and S9 B) and is consistent with a recent report on LAG-3 expression in DLBCL(38).

Immune cells in the TME can mediate chemoresistance through direct interaction with tumor cells or indirectly through interactions with other immune cells. Therefore, we sought to characterize the hyper-local microenvironment within 2-3 cell diameters (15µm) of REF enriched tumor cells (Figure 3D) by measuring its attraction of immune cell types. Attraction between cell types was defined as an observation of two cell types in closer proximity in the measured tissue than in randomly simulated tissue comprised of the same cell proportions (p<0.025). Comparing the tumor neighborhoods across treatment response, the REF tumor neighborhoods had significantly increased attractions with PD-L1^+^M2 macrophages (p=0.04831, t-test), however highly suppressive T_REG_, which were highly associated with REF at the case level, showed no significant enrichments in the hyper-local TME (p=0.74, t-test). In contrast, the hyper-local tumor neighborhoods associated with CR had uniformly increased CD4 phenotypes within the tumor environment compared to REF (p<0.05, t-test).

### Cross-cohort analysis shows differences in proportion and functional states of immune cell subsets between DLBCL and Hodgkin lymphoma

For patients who fail chemotherapy, treatment with antibodies against the PD-1/PD-L1 checkpoint have elicited clinical response rates close to 90% in patients with HL, while responses in patients with DLBCL are less than 10% (26,39). Therefore, we hypothesized that comparing the TME between reactive lymph nodes, HL, and DLBCL might reveal the basis for the difference in clinical responses to checkpoint inhibitors. We previously reported our analysis of the TME in 5 reactive lymph nodes and 22 cases of HL using a 36 marker IMC panel, of which 21 markers overlapped with the IMC DLBCL panel (Figure S10) (31). We integrated the TME analysis from both diseases after sub-setting each experiment to include only CD4, CD8, MAC, and T_REG_ cells, and identified the joint TME phenotypes (Figure 4A).

**Figure 4.**
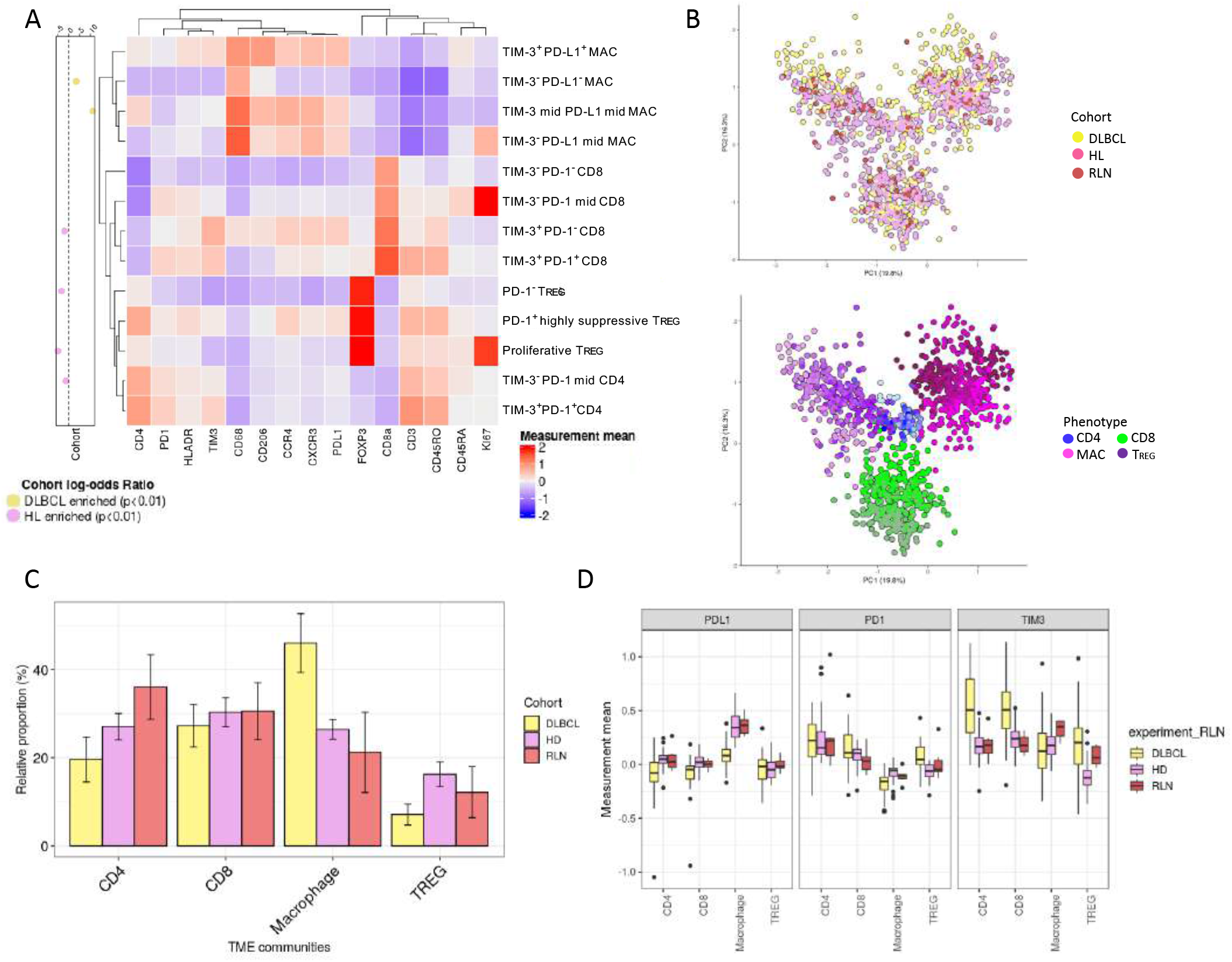
Cross-cohort analysis shows differences in proportion and functional states of immune sub-phenotypes between DLBCL and Hodgkin’s lymphoma (HL). A. The annotated heatmap depicting immune normalized expression of selected sub-phenotypes in both DLBCL and HL TME. Using the relative case proportions per sub-phenotype, the log-odds ratio (left) depicts the strength of association between the sub-phenotypes and DLBCL/HL (p<0.01). Complete heatmap of all sub-phenotypes Figure S12C B. After batch normalization, the PCA visually confirms the immune sub-phenotypes identified are well distributed across the two cohorts, indicating no cohort bias. Visual inspection confirmed using k-nearest neighbor batch effect test (kBET) (see Figure S11) C. Comparing relative immune proportions between DLBCL and HL shows that CD4 T cells (% difference=-10.2, 95% CI:(−16.2,-4.2)) and T_REG_ (% difference=-8.5, 95% CI:(−11.9,-5.02)) are significantly enriched in HL, while macrophages (% difference=22.1, 95% CI:(14.9,29.2)) are significantly enriched in DLBCL. CD8 were proportional across disease types (% difference=-3.4 95% CI:(−9.1,2.4)). D. Analyses of cell-state protein expression on each immune subset across the two cohorts show differences in functional states of immune subsets in DLBCL compared to HL. PD-L1 expression on macrophages is significantly higher in HL compared to DLBCL, whereas PD-1 expression is higher on CD8 and T_REG_ in DLBCL compared to HL. TIM-3 expression is significantly higher in DLBCL and is predominantly on CD4 and CD8 T cells.

After integrating both experiments, we tested for the presence of batch effects (Figure S11). By visual inspection of principal component analysis (Figures 4B, S12 and S13), the TME expression levels from both experiments were well inter-mixed. We quantitatively tested the level of inter-mixing between the two experiments using the K-nearest neighbor batch effect test and confirmed that the joint panel expression intensities on the TME were homogeneous in their expression (CD4 p=0.31, CD8 p=0.49, MAC p=0.43, T_REG_ p=0.08) (Figure S11) (40,41).

Comparing the TME phenotypes identified in DLBCL and HL, macrophages significantly increased by 24% in DLBCL compared to reactive lymph nodes (RLN) compared to the difference between HL and RLN (p<0.001) (Figure 4C), but within the respective HL/DLBCL populations of macrophages, global PD-L1 expression was lower in DLBCL (logFC= −0.26, BH q-value=7.35e-14) (Figure 4D, S14A). These results confirm the previously reported importance of PD-L1 on macrophages in HL (42).

Within macrophages, the relative proportion of TIM-3^+^PD-L1^+^ macrophage phenotypes were similar in both HL and DLBCL (Log-odds= −0.61, p=0.88), however the relative proportion of TIM-3 mid PD-L1 mid macrophages had the highest enrichment in DLBCL compared to HL (Log-odds=65, p<0.001), followed by TIM-3-PD-L1-macrophages (log-odds=2.1, p<0.001). CD4 cells were more abundant in HL compared to DLBCL, but the subset of TIM-3^+^T cells was increased in DLBCL (logFC=0.46, BH q-value<0.001) (Figures 4C and 4D). This corresponded with the TIM-3^−^PD-1^+^CD4 T cells significantly enriched in HL compared to DLBCL (Log-odds=0.38, p<0.001)

CD8 T cells were found in equal proportion in DLBCL and HL, however CD8 T cells had higher PD-1 (logFC=0.35, BH q-value=8.9e-09) and TIM-3 (logFC=0.44, BH q-value=2.3e-08) expression in DLBCL. Further, TIM-3^+^PD-1^−^ CD8 were significantly associated with HL (Log-odds=-1.7, p<0.001). Finally, T_REG_ cells were less abundant in DLBCL compared to HL (Bonferroni adjusted p-value=2.3e-05), but the proportion of the PD-1^+^T_REG_ subset was increased (logFC=0.23, BH q-value=7.3e-05) while the PD-1^−^Ki67^+^T_REG_ was significantly enriched in HL (Log-odds=-3.7, p<0.001) (Figure 4A,D, Figure S14 B).

### Spatial analyses reveal differences in tumor topology between GCB and non-GCB

The histopathology of DLBCL is predominantly characterized by large sheets of tumor cells without a distinct structure, however closer inspection reveals heterogeneity in the tumor cell and TME architecture. In order to characterize the single-cell topology we developed an algorithm for tumor cell-TME distance classification and tested it using synthetic images and B cells in normal lymph nodes. The distance classification algorithm we developed classifies a selected population into sub-clusters based on distances to neighboring cell types (Figure 5A). We constructed a demonstration set of four synthetic point patterns which simulated a lymph node (Figures S14 and S15) and successfully classified a point pattern in terms of the nearest neighbor distance (NND) to the other synthetic types. Next, we classified B cells in a normal lymph node based on their distance relationship to other cells. The topology analysis identified B cell spatial clusters that corresponded to regions in the light and dark zone of germinal centers, the mantle zone of the follicle, and T cell rich paracortex regions (Figures S15A and S15C).

**Figure 5.**
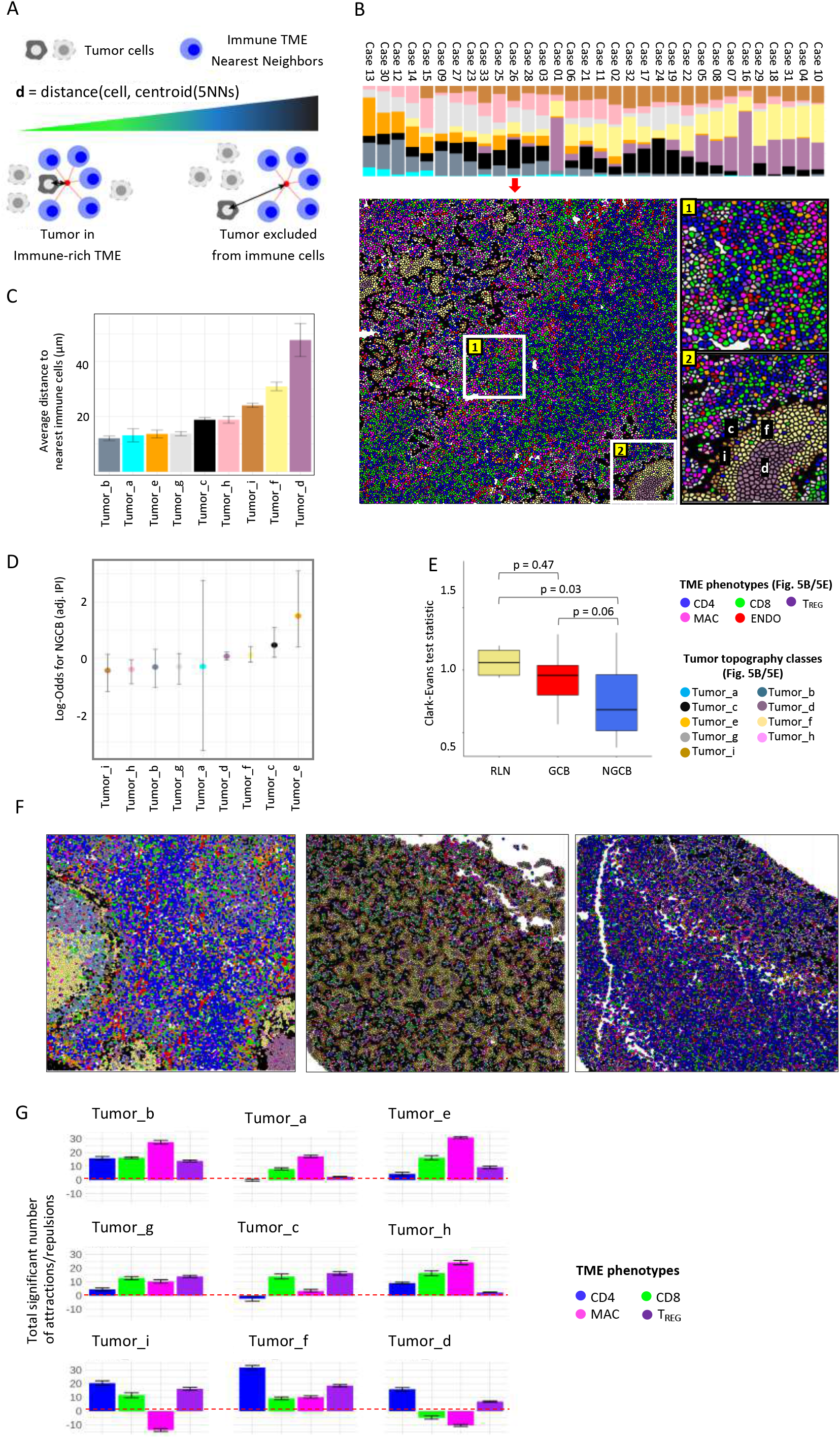
Spatial clustering reveals differences in tumor topology that associate with COO and TME abundance. A. Neighborhood analysis of cells describes the local arrangement of cells in the tumor. The metric is calculated for each cell by locating the 5 nearest cells belonging to the immune TME, locating the centroid of those nearest neighbors, and measuring the distance from the centroid to the original cell. A smaller distance metric indicates that a cell is embedded within the immune TME, a longer distance denotes exclusion of tumor cells from immune cells. B. Nine classes of tumor topology were identified and were well distributed across cases, ordered from immune-cold to immune-hot. Intra-tumor spatial heterogeneity is depicted in a representative annotated image from case 26. The first inset shows tumor classes that are more intermixed with the immune cells. The second inset shows tumor spatial arrangement structures similar to the geological topography, with tumor_d situated at the “core”, tumor_f at the inner “mantle”, tumor_i at the outer “mantle”, and tumor_c at the “crust” of tumor clusters. C. Histogram showing the ordered average distance from each tumor topology class to its nearest immune cells. Tumor topology classes were ordered by their distance/proximity to the TME. D. Estimates from a multivariate logistic regression model adjusted for IPI, the log-odds of NGCB vs. GCB for all tumor topology class abundances are depicted. Holding all the other terms constant, tumor_e (log-odds=1.498, p=0.026) is significantly enriched in NGCB, where tumor_c (log-odds=0.46,p=0.08) and tumor_h (log-odds= −0.4, p=0.057) had marginal associations with NGCB and GCB respectively. E. Clark-Evans aggregation index quantifies the level of spatial regularity (index>1), or clustering (index<1), and was applied to the B-cell topology classes in reactive lymph node (RLN) and DLBCL (GCB and NGCB). The Clark-Evans index (box plots) indicated the B-cell topology classes in non-GCB cases were significantly more clustered compared to RLN (Tukey p=0.03), whereas GCB had spatial regularity similar to RLN (Tukey p=0.47). There was marginal difference comparing GCB to NGCB (Tukey p=0.058). This indicates that the spatial distribution of the malignant B cells in GCB tumors more closely resembles the B-cell architecture of normal follicles, while malignant B cell in ABC tumors were more dispersed. F. Representative ROIs of RLN (Left, Clark-Evans Index =1.16), GCB (Middle, Clark-Evans Index=1.29), and NGCB (Right, Clark-Evans Index=0.54). G. From the tumor zone reference, the total significant attractions/repulsions of each major cell component were summed in each tumor topology (with 95% confidence interval). Positive values indicate attraction while negative values indicate significant repulsions. The CD4 were enriched within tumor core and mantle neighborhoods, while CD8 were depleted in the core and mantle regions but enriched in the crust and dispersed regions. Macrophages were found in the dispersed, crust and mantle areas but were depleted from the core. The x-axis denotes the major TME, the y-axis denotes the total sum of the significant signed interactions (p<0.05). The tumor zones are ordered by distance to TME from closest (tumor b) to furthest (tumor d).

Having validated the tumor-TME distance clustering approach on synthetic images and normal lymph node, we applied our distance classification algorithm to the tumor cells in DLBCL and identified nine spatial clusters, or tumor neighborhoods, based on tumor distance to the TME. The rarest topology was class “a”, or “tumor_a”, (0.93% prevalent), whereas tumor_f (21.1%), tumor_d (15.3%) and tumor_c (15.2%) were most common (Figure 5B). The topology classes had significant heterogeneity across the immune proportions (p=0.01507; ANOVA) which is not unexpected since abundance of immune cells impacts their distance to tumor cells in a given tissue. From visual inspection it was clear that some tumor cells were dispersed among immune cells such as tumor_b and tumor_e (Figure 5B, inset 1), while other tumor cells formed tight layered clusters such as tumor_d, tumor_f, and tumor_i (Figure 5B, inset 2)

Considering that tumors are three-dimensional, we grouped the nine tumor spatial clusters into four groups inspired by the geologic structure of Earth: core, mantle, crust and dispersed. Tumor_d (average centroid distance to immune (dist)= 47.9µm) was defined as the tumor “core” because it was characterized by the furthest distance to the tumor-immune interface. Tumor_f (dist= 30.9µm) followed by tumor_i (dist= 24.1µm) were the next clusters in order of distance to tumor-immune interface and were defined as the tumor “mantle” because they were consistently adjacent to the core (Figure 5B, inset 2). Tumor_c (dist= 18.9µm) and tumor_h (dist= 18.9µm) represented the boundary to the tumor-immune interface, or “crust” because it was adjacent to both the TME interface and mantle/core regions. Tumor_g (dist= 13.8µm), tumor_e (dist= 13.7µm) tumor_a (dist= 13.2µm), and tumor_b (dist= 12.2µm) had the shortest distances to the TME and formed unorganized tumor clusters (Figure 5B, inset 1). Hence, we defined these scattered cells as “dispersed” regions of the tumor. Using this conceptual framework, we were able to spatially organize and interpret the tumor spatial clusters in terms of proximity to the TME.

The nine tumor spatial clusters were tested for association with COO using a multivariate logistic regression model. After adjusting for IPI, tumor_e (p=0.026), was significantly associated with non-GCB (Figures 5D, Figure S16A). Tumor_e represented scattered tumor cells in regions rich with lymphocytes (Figure 5B, inset 1). Tumor_c, found at the tumor immune interface was marginally associated (log-odds=0.46, p=0.08) with non-GCB (Figure 5B, inset 2). These results are consistent with non-GCB tumors having increased interactions with immune cells.

To further explore if COO was associated with differences in tumor topology, we quantified the level of spatial clustering across each case, using the Clark-Evans aggregation index. The Clark-Evans aggregation index has been used in fluorescence microscopy studies and geographical research as a measure of spatial organization (43,44). Here, we used the standardized Clark-Evans index to compare the B cells in normal lymph node to the malignant B cells in GCB and non-GCB cases (Figure 5E-F). The malignant B cells in GCB patients had similar spatial ordering to that of B cells in RLN (Tukey adjusted p-value=0.47), whereas the malignant B cells in non-GCB had significantly different spatial organization (Tukey adjusted p-value=0.03). Comparing GCB to non-GCB, there was marginal difference in their spatial distribution (Tukey adjusted p-value =0.058). Further, as the spatial patterns of tumor cells become less clustered, the risk of dying decreased (Cox hazard estimate=-1.89, 95% CI: (−4.52, 0.75)). Increasing architectural disorganization is routinely recognized in carcinoma as a feature of tumor aggressiveness but is not routinely described in DLBCL histopathology. Our results quantify this feature and demonstrate that decreasing spatial regularity in DLBCL architecture correlates with both COO and survival.

We next explored whether different immune subsets had differential interactions with the tumor spatial clusters. For each of the major immune classes we determined significant spatial attractions or repulsions towards each tumor spatial cluster (Figure 5G). We observed that CD4 T cells had significant associations within the tumor mantle/core compared to the crust/dispersed regions (OR=6.38, 95% CI: (2.91,14.43), Fisher’s exact p value=3.9e-06). This suggests that CD4 T cells were more likely to penetrate into tumor regions furthest from the TME interface. Whereas, the CD8 T cells had decreased association with mantle/core compared to the crust/dispersed (OR=0.16, 95% CI: (0.05,0.41), Fisher’s exact p value=5.6e-05), which suggests that CD8 are restricted to outer regions of the tumor cell-immune interface.

Macrophages were able to partially penetrate into the mantle areas but were depleted from deeper core areas (tumor_d) and had marginal decreased association at the tumor mantle/core compared to the crust/dispersed (OR=0.21, 95% CI: (8.5e-03, 1.1), Fisher’s exact p value=0.12). Encouraged by these preliminary results, we next sought to comprehensively characterize the immune cells in each tumor zone.

### DLBCL comprehensive spatial interactions between TME and tumor topology, functional state profile, mutations, and patient clinical association

To comprehensively characterize the topology of tumor immune interaction, we adapted a previously described method for examining spatial interaction in IMC data to look for spatial interactions between tumor spatial clusters and immune phenotype sub-clusters within a 15µm interaction zone(35). Figure 6A summarizes the spatial interactions between immune phenotypes and tumor spatial clusters, the protein expressions on each cluster, as well as their multivariate association (IPI adjusted) with genetic mutations and Hans COO. For example, tumor_e, which was enriched in C4 DLBCL genetic subtype (p<0.05) subtype and was classified as a dispersed tumor type, had enriched interactions with activated and exhausted CD8 T cells, and Memory CD4 T cells. Tumor_c, found at the tumor immune interface, showed significant negative association with C2 (p<0.05), and was not associated with the C5 genetic subtype, and was significantly associated with participants having CD79b mutation (Benjamini-Hochberg adj. P-value=0.02). Tumor_c was enriched for interactions with Ki67^+^ memory CD4, TIM-3^+^PD-1^+^LAG-3^+^ exhausted CD8 T cells, and highly suppressive T_REG_.

**Figure 6.**
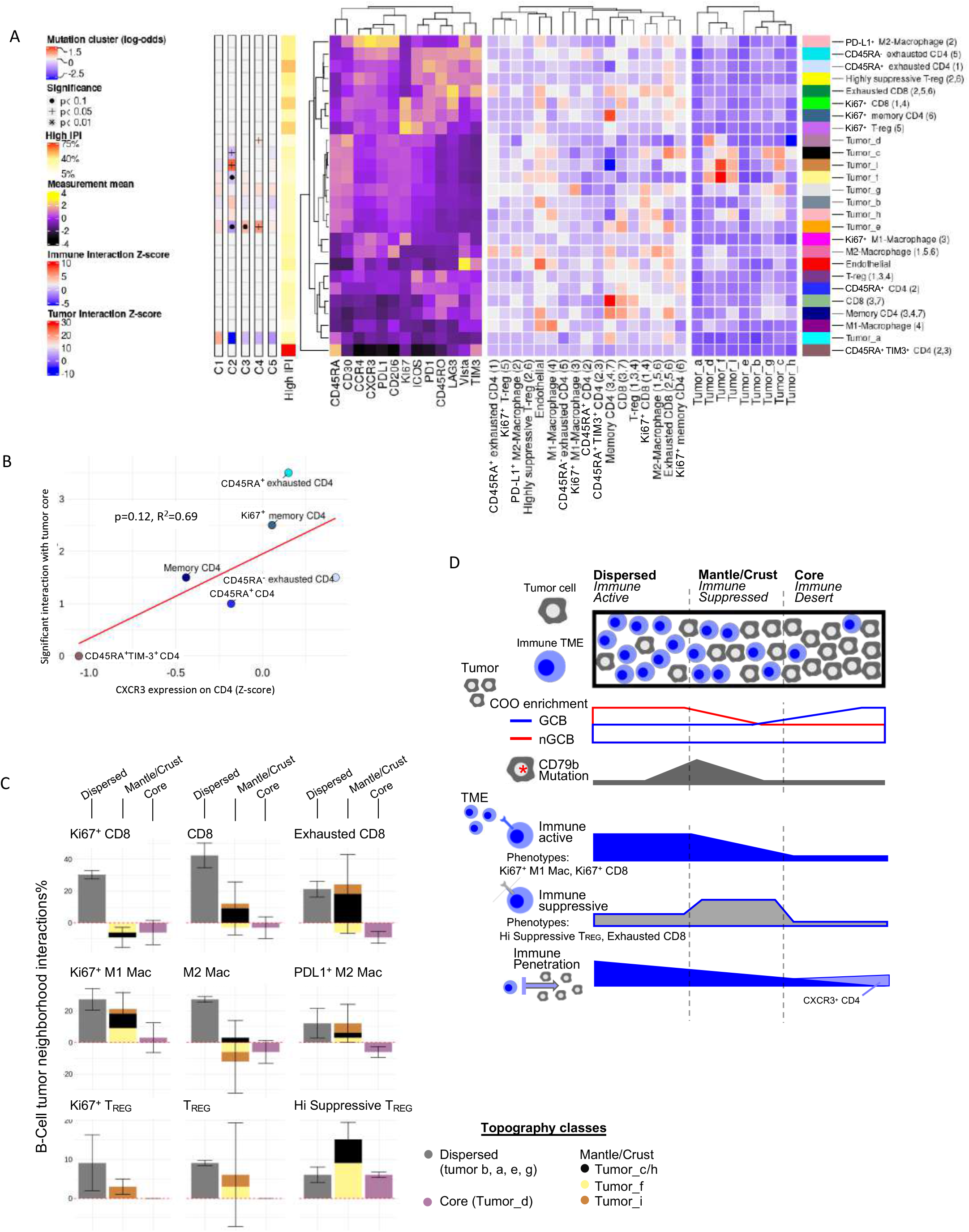

Tumor_f, found in the inner mantle was enriched for endothelial cells, and depleted for CD8 subsets. This region was enriched for both highly suppressive T_REG_ as well as M1-macrophages and this heterogeneity may reflect the trafficking of immune cells from the peripheral blood. Tumor_d, which defined the core regions, contained a sparse TME neighborhood with depletion of most immune subsets with the exception of occasional CXCR3^+^CD45RA^+^CD4 and highly suppressive T_REG_ cells. Tumor_d regions were significantly associated with the C4 genetic subtype (p<0.05) and were not associated with refractoriness to chemotherapy, however the presence of these pockets of immune deserts may explain the lack of efficacy of checkpoint inhibitors in DLBCL and could have implications for cellular therapies such as CAR-T cells. While penetrating CD4 cells were overall rare, there was a moderate correspondence between CD4 and CXCR3 co-expression, and the total number of significant spatial interactions with the tumor core (p=0.12, R^2^=0.69, Figure 6B). This suggests that CXCR3 plays a role in granting CD4 cells entry into otherwise restricted tumor sites and suggests a possible target for enhancing immune cell penetration into the tumor core regions.

To summarize our findings, we grouped the nine tumor spatial clusters into three zones: dispersed (tumor_b, tumor_a, tumor_e, and tumor_g), crust/mantle (tumor_c, tumor_h, tumor_f, and tumor_i) and core (tumor_d) and examined the immune cells surrounding tumor cells in each zone (Figure 6C). The dispersed zone showed enrichment for proliferative subsets of CD8, macrophage, and T_REG_ surrounding tumor cells, which was consistent with high immune activity. The mantle/crust zone was most notable for enrichment of exhausted CD8 and highly suppressive T_REG_ around tumor cells suggesting a zone of immune suppression. The core region was depleted for most immune phenotypes consistent with an immune desert. These three zones of immune activation, immune suppression, and immune desert corresponded to the tumor spatial zones of dispersed, mantle/crust and core, respectively (Figure 6D). On to this topology we overlaid COO and mutation status. Finally, by creating an ordered topology from high to low immune density we were able to identify CXCR3^+^CD4 cells as having the most tumor penetrating properties.

## DISCUSSION

In this highly multiplexed single-cell study of the tumor immune architecture in DLBCL, we have quantified the intra-tumor heterogeneity of B cell lymphoma architecture and identified immune phenotypes that correlate with COO, tumor mutations, and response to therapy. A similar effort in breast cancer was recently reported, however this is the first such report in lymphoma where the high cellular density and the co-expression of markers on tumor and immune cells make this type of study particularly challenging (13,14).

Within DLBCL, a recent large-scale multiplexed study with a focus on the role of PD-L1 and PD-1 on CD20^+^ DLBCL cells was performed using immunofluorescence, but that study was limited to 13 markers(37). In that study, Xu-Monette et al. identified that PD-1^+^CD8 cells were associated with poorer overall survival. PD-1 expression on CD4 also showed adverse prognosis on univariate analysis at low cut offs, but a paradoxical better prognosis when a high level cutoff for PD-1 expression was used. These results suggest that there may be sub-populations within the PD-1^+^CD4 population that have differential effects on survival. Indeed, in our study we observed that CD4 and CD8 T cells with PD-1 expression could be further divided into activated Ki67^+^T cells and terminally exhausted PD-1^+^TIM-3^+^LAG-3^+^ triple positive T cells, highlighting the importance of highly multiplexed analysis.

Furthermore, we demonstrated that subgroups enriched in refractory patients showed higher PD-L1 and PD-1 levels compared to CR, which is consistent with previous results. PD-1/PD-L1 expression differences are possibly coupled to spatial expression patterns. The co-localization of PD-L1^+^M2 macrophages with tumor suggests that these cells mediate chemoresistance through direct interactions with tumor cells, while highly immunosuppressive T_REG_, which are common in REF patients but not tumor-adjacent, exert their effects primarily through interactions with other immune cells. The ranges of influence that immune cells in the TME can have in modulating chemoresistance of malignant B cells is a topic for future studies.

Our multivariate analysis of tumor spatial clusters identified that exhausted CD8 T cells were excluded from tumor dense mantle and core regions while selected PD-1^+^CD4 T cells were better able to access the tumor core regions. This arrangement is somewhat reminiscent of the architecture of a germinal center with CD8 T cells relegated to the surrounding T cell zone and T follicular helper cells (PD-1^+^CD4) interacting with proliferating B cells in the center. In contrast to the normal GC, however, we also saw T_REG_ cells penetrating the tumor core regions which could provide an alternative explanation for the lack of infiltrating CD8 cells. Loss of MHC Class I expression has been previously reported in DLBCL (49%-61%) and could also contribute to a lack of CD8 T cell engagement with the tumor cells (45).

Our observation that tumor core areas, which exclude immune cells, are found in most DLBCL cases may have important implications for clinical failure of ICI therapies and resistance to cellular therapies such as chimeric antigen receptor (CAR) T cells. Among the CD4 cells, we observed that CXCR3-high subsets, while rare, had infiltrating potential in lymphoma, similar to previous reports in other diseases (46–48). Importantly, Xu-Monette et al. reported that up-regulation of *CXCL9*, a ligand for CXCR3, was found in DLBCL with higher T cell infiltration, further supporting the role of CXCR3 in T cell homing to tumor core regions(37). We hypothesize that activating CXCR3 or enforcing expression of CXCR3 on CAR-T might improve immune cell penetration into lymphoma core regions. Applications of IMC analysis on treatment biopsies after CAR-T therapy could be particularly helpful to understanding treatment response and failure in this context.

Given the clinical failure of PD-1/PD-L1 inhibitors in DLBCL, identifying novel immuno-oncology targets for DLBCL remains critically important. DLBCL checkpoint therapy non-responsiveness can be further understood by investigating alternative check point molecules beyond PD-1/PD-L1 such as TIM-3, LAG3, and/or VISTA. Interestingly, DLBCL had higher levels of PD-1 on T cells compared to HL, an observation that is consistent with PD-1 being a poor biomarker of checkpoint response. Additionally, the comparison between DLBCL to HL identified TIM-3 as over-expressed primarily in CD4 and CD8 T cells which highlights it as a therapeutic target (49). Furthermore, PD-1 was enriched in T_REG_ in DLBCL, whereas TIM-3 on T_REG_ was not, which demonstrates the importance of identifying therapeutic targets that are differentially enriched on specific CD4/CD8 immune subsets. Given the high level of PD-1 on T_REG_ that we observed, anti-PD-1 antibodies may have led to increased activity of T_REG_ resulting in paradoxical suppression of immune response after check point therapy and clinical failure. These observations demonstrate that comparisons of TME between ICI responsive and non-responsive sub-types of lymphoma can suggest potential mechanisms of ICI sensitivity and resistance.

In order to perform our comparisons between DLBCL and Hodgkin lymphoma, we integrated IMC data from experiments performed using different antibody panels. The increased dynamic range of signal with IMC, compared to IHC, creates challenges with increased variability in signal intensity across experiments. Here, using approaches to normalize and standardize the data, we were able to demonstrate meaningful comparisons between different data sets. There were limitations to this method, however, and even after data integration several markers had to be excluded because of poor normalization. As additional studies using imaging mass cytometry or other similar high multiplexed single cell imaging techniques are published, methods to measure uncertainty across experimental procedures will need to be developed.

One potential limitation of our study is that we only examine a relatively small part of the tumor area (approximately 1mm^2^) and may not have captured biologically relevant heterogeneity outside the region of interest. However, within that small area, we are able to discern a high degree of spatial and functional heterogeneity and describe a complex tissue architecture of immune response and suppression that corresponds to clinical feature. Optimal sample size will also depend on the specific tumor type and will need to be determined empirically. In our data, seven cases had at least one duplicate core on the TMA. Analysis by PCA suggested that most duplicates clustered together (Figure S1C). This would suggest that small sample areas may be adequate to understand the complexity of tumor immune responses, at least in DLBCL. This is important because on-treatment and progression biopsies, which are critical to understand the kinetics of immune response and failure of immune-oncology agents, are often obtained as core needle biopsies, which are much smaller than diagnostic tissue samples.

Another limitation of these data is that we only study diagnostic samples of patients prior to chemotherapy and not prior to treatment with immune-oncology agents. Since DLBCL responses to ICI are rare, we compared the TME in DLBCL to Hodgkin’s lymphoma, where most patients achieve clinical responses. However, there are many differences between HL and DLBCL that could also account for the difference in TME, and the agreement between the response to ICI in these two tumor types and our observations would need to be experimentally verified. Further validation of additional patient data using IMC would reinforce these findings and determine if the results reported in this study that lie at the border of significance are truly significant.

Finally, the multiplex analysis provided by IMC could help guide the next generation of combination ICI therapies, including newer agents targeting CCR4, LAG-3, and TIM-3, or novel cellular therapies and bi-specific antibodies currently in development for lymphoma. Combining IMC with multi-omics profiling of pre-treatment and on-treatment biopsies will be powerful translational tools for understanding mechanisms of resistance and clinical failure for ICI and cellular therapies. In summary, IMC analysis of lymphoma reveals phenotypic and spatial structure in the TME that gives new insights to tumor immunology.

## Data Availability

data is public available, and can be used/shared

https://github.com/arcolombo/singleCell_DLBCL

## STAR Methods

### KEY RESOURCES TABLES

| REAGENT or RESOURCE | SOURCE | IDENTIFIER |
| --- | --- | --- |
| **Antibodies** |  |  |
| BCL-2 (clone EPR17509) | Fluidigm | Cat# 3146019D; RRID:<br>AB_2811012 |
| BCL-6 (clone K112-91) | Fluidigm | Cat# 3147020D; RRID:<br>AB_2811013 |
| CD134 (polyclonal) | Fluidigm | Cat# 3151024D; RRID:<br>AB_2811014 |
| CD183 (clone G025H7) | Biolegend | Cat# 353733; RRID:<br>AB_2563724 |
| CD194 (clone 205410) | Fluidigm | Cat# 3149003A; RRID:<br>AB_2811015 |
| CD20 (clone H1) | Fluidigm | Cat# 3161029D; RRID:<br>AB_2811016 |
| CD206 (clone 5C11) | Fluidigm | Cat# 3999999-2 |
| CD3 (Polyclonal) | Fluidigm | Cat# 3170019D; RRID:<br>AB_2811048 |
| CD31 (clone C31.3) | Abcam | Cat# ab212709; RRID:<br>AB_2811049 |
| CD34 (clone QBEnd/10) | Abcam | Cat# ab213054; RRID:<br>AB_2811050 |
| CD4 (clone EPR6855) | Fluidigm | Cat# 3156033D; RRID:<br>AB_2811051 |
| CD45RA (clone HI100) | Fluidigm | Cat# 3155011B; RRID:<br>AB_2810246 |
| CD45RO (clone UCHL1) | Fluidigm | Cat# 3173016D; RRID:<br>AB_2811052 |
| CD68 (clone KP1) | Fluidigm | Cat# 3159035D; RRID:<br>AB_2810859 |
| CD8α | Fluidigm | Cat# 3162034D; RRID:<br>AB_2811053 |
| c-Myc p67 (clone 9E10) | Fluidigm | Cat# 3164025D; RRID:<br>AB_2811054 |
| Ephrin B2 | Abcam | Cat# ab233246; RRID:<br>AB_2811055 |
| FoxP3 (clone 236A/E7) | Thermo Fischer | Cat# 14-4777-82; RRID:<br>AB_467556 |
| Granzyme B (clone<br>EPR20129-217) | Fluidigm | Cat# 3167021D; RRID:<br>AB_2811057 |
| Histone 3 (clone D1H2) | Fluidigm | Cat# 3176023D; RRID:<br>AB_2811058 |
| HLA-DR (clone YE2/36 HLK) | Fluidigm | Cat# 3174023D; RRID:<br>AB_2811059 |
| ICOS (clone D1K2T) | Fluidigm | Cat# 3148021D; RRID:<br>AB_2811060 |
| Ki67 (clone B56) | Fluidigm | Cat# 3168022D; RRID:<br>AB_2811061 |
| LAG-3 (D2G40) | Fluidigm | Cat# 3153028D; RRID:<br>AB_2811062 |
| PD-1 (clone NAT105) | Biolegend | Cat# 367402; RRID:<br>AB_2565782 |
| PD-L1 (clone 28-8) | Abcam | Cat# ab209889; RRID:<br>AB_2811063 |
| PD-L2 (clone 176611) | Fluidigm | Cat# 3172028D; RRID:<br>AB_2811066 |
| pStat3 [Y705] (clone 4/P-Stat3) | Fluidigm | Cat# 3158005A; RRID:<br>AB_2661827 |
| Tbet (clone D6N8B) | Fluidigm | Cat# 3145015D; RRID:<br>AB_2811067 |
| TIM-3 (clone D5D5R) | Fluidigm | Cat# 3154024D; RRID:<br>AB_2811068 |
| Vimentin (clone RV202) | Fluidigm | Cat# 3143029D; RRID:<br>AB_2811069 |
| Vista (clone D1L2G) | Fluidigm | Cat# 3160025D; RRID:<br>AB_2811070 |
| **Metal Isotopes** |  |  |
| 142Nd | Fluidigm | Cat# 201142A |
| 143Nd | Fluidigm | Cat# 201143A |
| 144Nd | Fluidigm | Cat# 201144A |
| 145Nd | Fluidigm | Cat# 201145A |
| 146Nd | Fluidigm | Cat# 201146A |
| 147Sm | Fluidigm | Cat# 201147A |
| 148Nd | Fluidigm | Cat# 201148A |
| 149Sm | Fluidigm | Cat# 201149A |
| 150Nd | Fluidigm | Cat# 201150A |
| 151Eu | Fluidigm | Cat# 201151A |
| 152Sm | Fluidigm | Cat# 201152A |
| 153Eu | Fluidigm | Cat# 201153A |
| 154Sm | Fluidigm | Cat# 201154A |
| 155Gd | Fluidigm | Cat# 201155A |
| 156Gd | Fluidigm | Cat# 201156A |
| 158Gd | Fluidigm | Cat# 201158A |
| 159Tb | Fluidigm | Cat# 201159A |
| 160Gd | Fluidigm | Cat# 201160A |
| 161Gd | Fluidigm | Cat# 201161A |
| 162Dy | Fluidigm | Cat# 201162A |
| 163Dy | Fluidigm | Cat# 201163A |
| 164Dy | Fluidigm | Cat# 201164A |
| 166Er | Fluidigm | Cat# 201166A |
| 167Er | Fluidigm | Cat# 201167A |
| 168Er | Fluidigm | Cat# 201168A |
| 169Tm | Fluidigm | Cat# 201169A |
| 170Er | Fluidigm | Cat# 201170A |
| 172Yb | Fluidigm | Cat# 201172A |
| 172Yb | Fluidigm | Cat# 201142A |
| 173Yb | Fluidigm | Cat# 201173A |
| 174Yb | Fluidigm | Cat# 201174A |
| 175Lu | Fluidigm | Cat# 201175A |
| 176Yb | Fluidigm | Cat# 201176A |
| **Biological Samples** |  |  |
| SignalSlide® PD-L1 control | Cell Signaling Technology | Cat# 13747 |
| **Chemicals, Peptides and Recombinant proteins** |  |  |
| m-Xylene anhydrous, >99% | Sigma Aldrich | Cat# 296325-2L |
| Ethanol | Sigma Aldrich | Cat# 459836-2L |
| PBS (Calcium & Magnesium free, pH7.4) | Genesee Scientific | Cat# 25-507 |
| Trizma base | Sigma Aldrich | Cat# T1503-250G |
| EDTA | Sigma Aldrich | Cat# E1644-250G |
| Tween 20 | Fischer Scientific | Cat# BP337-100 |
| BSA (DNase and Protease-free Powder) | Fischer Scientific | Cat# BP9706100 |
| Triton X-100 | Sigma Aldrich | Cat# T8787-100ML |
| Cell ID Intercalator (Iridium) | Fluidigm | Cat# 201192A |
| **Software and Algorithms** |  |  |
| CellProfiler (version 2.1.0) |  | https://cellprofiler.org/previous_releases;<br>RRID:SCR_007358 |
| Ilastik (version 1.3.0) |  | https://www.ilastik.org/download.html; RRID:SCR_015246 |
| Rstudio (version 3.6.1) |  | http://www.rstudio.com;<br>RRID:SCR_000432 |

### LEAD CONTACT AND MATERIALS AVAILABILITY

Further information and requests for raw data, codes, and other resources should be directed to the Lead Contact, Akil Merchant.

### SUBJECT DETAILS

This retrospective study included a subset of 33 patients from a previously studied cohort of 85 patients diagnosed with de novo DLBCL at Los Angeles County and University of Southern California (USC) Medical Centers between 2002 and 2012 (32). The sub cohort was representative of the primary cohort and was not selected other than looking for samples with adequate remaining tissues for further analyses (Figure S1). This study was approved by the USC Health Sciences Institutional Review Board.

## METHODS DETAILS

### Tissue Microarray and Immunohistochemistry

Three of the six tissue microarray (TMA) blocks from the parent cohort with sufficient remaining tissues representative of viable tumor were obtained from the Pathology archive of Los Angeles County and USC Medical Centers. Immunohistochemistry (IHC) staining was performed on 4-μm tissue sections using DAKO ready-to-use antibodies with the EnVision FLEX and FLEX+ visualization systems (DAKO, Glostrup, Denmark) on an automated immunostainer (Autostainer Link 48, DAKO). Detailed IHC staining protocol and scoring methods have previously been published (32). TMA cores were 2mm in diameter.

### Imaging Mass Cytometry staining

Three of the six TMAs with optimal quality of remaining tumor tissues from the larger cohort study were selected for this study. The TMAs contained 42 cores of FFPE DLBCL tissues from 33 patients and 2 cores from liver tissues. FFPE sections of 4-µm were baked at 60°C for 90 minutes on a hot plate, de-waxed for 20 minutes in xylene and rehydrated in a graded series of alcohol (100%, 95%, 80% and 70%) for 5 minutes each. Heat-induced antigen retrieval was conducted on a hot plate at 95°C in Tris-EDTA buffer at pH 9 for 30 minutes. After blocking with 3% BSA in PBS for 45 minutes, the sections were incubated overnight at 4°C with a cocktail of 32 antibodies tagged with rare lanthanide isotopes obtained from Fluidigm (Table S1). Titration for PD-1 antibody was performed on tonsil tissue (follicular T helper cells in the germinal center). PD-L1 titration was done on a commercial slide containing formalin-fixed paraffin-embedded cell pallets of HDLM-2 (PD-L1+) and PC-3 (PD-L1-) cell lines from Cell Signaling Technology (Key resources table). Validation and titration of all other markers were done on control tonsil tissue (Figure S3). HLA-DR, pSTAT3, PD-L2 and Ephrin-B2 antibodies showed unspecific staining patterns in the control tonsil tissue and therefore were excluded from analyses.

### Tissue imaging and ablation

All cores were evaluated by two pathologists (I.S. and M.H.) to identify region of interest (ROI) on H&E. Slides were analyzed using the Fluidigm Hyperion Tissue Imager system that couples laser ablation with mass spectrometry (50). Laser beam of 1-µm^2^ spot size was used to ablate tissue area of 1000-µm^2^ per core at a frequency of 200 Hz. The metal isotopes were simultaneously measured and indexed against the location of each spot to generate intensities and digital spatial maps of the ablated tissues. Detailed descriptions of the ablation techniques have been previously described (30,51).

### Image Analysis Pipeline

The ion counts for each metal-labeled antibody and slide location were compensated for the cross talk between channels then converted to OME-TIFF images (52,53). Images for each antibody were scaled using the 95 percentiles of the cumulative signal to remove hot spot pixels and normalized across acquisitions (34).

### Image Segmentation

Channels representing distinct morphological features for cell nuclei (i.e. Ir193-DNA Intercalator, Histone H3, foxP3, Ki67) and membrane staining (i.e. CD8, CD68, CD45RA) were used for the Ilastik pixel classification training to predict nuclei, membrane/cytoplasm and background pixel class using cropped 2x scaled images (54). The probability maps were segmented using CellProfiler by subtracting the membrane probability map from the nuclei and then expanding the nuclei by 4 pixels (34).

## QUANTIFICATION AND STATISTICAL ANALYSES

### Data transformation and normalization

The presented data used normalization such as the hyperbolic-arc-sine transformation, and Min/Max normalization at the 99^th^ percentile (14,36). The authors for Phenograph and t-SNE recommended using the 99^th^ percentile to clip the data to [0,1] scale and remove outliers, which we followed. For the cluster expression estimates, and spatial expression heterogeneity modeling, we used the mixed-effects linear model with the single-cell marker expression intensities scaled to a standard normal distribution across each ROI. Thus, for linear modeling we ensure that for all tissues, each marker expression was on the standardized Gaussian distribution. For Cox proportional hazards estimates, the relative proportions of each cluster were used as features, with survival times (N=30, events=7) in univariate models.

### Analysis workflow

The exploratory analysis used histoCAT, and downstream tSNE clustering using ‘Rtsne (v.0.15)’,’lme4 (v.1.1.21)’ R (v.3.6.3) packages for clustering and mixed-effects linear models (35). The neighborhood analysis used Bodemiller repository (https://github.com/BodenmillerGroup/neighbouRhood).

All the figures and codes for the analysis of this project (figure 1-6) are available here: https://github.com/arcolombo/singleCell_DLBCL

### Clustering and metaclustering

#### Phenotypic clustering

The images along with the masks were imported into histoCAT software for initial evaluation. Cell features were extracted and imported into R statistical software environment (35). We hierarchically performed meta-clustering to identify cell “phenotypes”. The first step under-clustered the data using lineage related markers (BCL2, BCL6, CD20, CD206, CD3, CD30, CD31, CD4, CD45RA/RO, CD68, CD8, EphrinB2, FOXP3 and HLADR) clustering each ROI (nearest neighbor, k_1_=45) and then clustering on the centroids (nearest neighbor, k_2_=15) (36). The major cell classification per case identified 14 meta-clusters, and each ROI had on average 1,145.174 cells per meta-cluster. Quality control analysis performed Phenograph (k=50) on each major cell component separately to ensure that the major cell expression was homogeneous on the corresponding marker. Re-assignments of CD4 comprised of 1.54%, the initial tumor component reassigned 6.21% to CD8, CD4, MAC, and endothelial classes. Following the re-assignments, we identified minor clusters for each major cell component by re-performing meta-clustering (k_1_=45, k_2_=15) on each TME component separately, with the inclusion of inducible state markers (cMYC, CCR4, CXCR3, Granzyme, ICOS, Ki67, LAG-3, PD-1, PD-L1, PD-L2, TBET, TIM-3, Vimentin, VISTA, and pSTAT3) and morphological features (Area, eccentricity, solidity, perimeter, percent touching, number of neighbors).

### TMA and replicate divergence analysis

The cohort comprised of 3 TMAs, and principal component analysis (PCA) was used to determine the presence of batch effects across TMAs. After annotating the 14 meta-clusters into major cell components, we performed PCA on the relative proportions per ROI and the PCA showed well-mixed visual representation of the variability of the data that did not have any TMA specific grouping. Case 17, 18, 21, 26, 27 and 31 had replicate ROIs taken and case 30 had triplicate ROIs. Using the R package ‘entropy (v.1.2.1)’, we computed the Kullback-Leibler (KL) divergence using a given replicate ROI relative frequencies per major cell component and compared a given replicate to the case relative average of the major components. The KL divergence scores per ROI were all below 0.10, however Case 26 had increased ROI heterogeneity (KL divergence of 0.26). The PCA analysis of the replicates showed minimal distance separations with the exception of Case 26. The replicates were mostly similar, with the exception of Case 26, and for the analysis of patient clinical variables we used the case averages across the ROIs.

### Association between genetic mutations, cell of origin and proportions of tumor-immune sub-phenotypes

The cluster analysis followed the guidelines from the authors, which used the 99^th^ percentile normalization to remove outliers by scaling to relative 99^th^ percentile of each marker. The heatmap visualization standardized each ROI to a standard normal distribution. The heatmap depicts the mean normalized intensities, which ensures that the tissues were standardized. The clustering of all the phenotypes present used bootstrapping (100) using ‘pvclust (v.2.2.0)’ and hierarchically clustered using sub-cluster means (Euclidean distance, Ward’s method) ‘dendsort (v.0.3.3)’ R package.

### Clinical association of sub-clusters to COO and mutation

In order to test and measure the odds ratio between sub-clusters and HANS cell of origin classification of GCB and NGBC, we constructed a 2×2 contingency table at the patient/case level. The relative proportions of a given sub-cluster present/absent in each patient stratified on the HANS classification formed the 2×2 contingency table. The simple non-parametric odds ratio of association between the sub-clusters and HANS are depicted in Figure 2 and the p-values used the Pearson’s Chi-square test of association. The HANS odds ratio depicted if the OR is greater than 1, or less than 0.90 with p<0.01. The odds ratio indicates the strength of association between a given cluster and GCB/NGBC classification.

The international prognostic index scores [0-5] were measured, and a median cutoff (>3) was used to identify patients with high IPI scores. For each sub-cluster the relative proportion of high IPI patients were computed providing a simple proportion of the relative proportion of high IPI subjects present within a sub-cluster.

BCL2 and MYC protein were measured by immune-histochemistry. BCL2 overexpression was measured using a 40% cutoff judged by IHC, and MYC over expression was determined as 70% threshold. Patients classified as double expressors were identified as BCL2 above the 40% and MYC above the 70% threshold. A mixed-effect linear regression model was used to identify significantly above average expression of cMYC and BCL2 or cMYC and BCL6 (α-level =0.05). Each phenotype subset was compared with the average expression for the corresponding major phenotype and significantly above average expression of cMYC and BCL2 or cMYC and BCL6 were tested (α-level =0.05). r. Tumor_6 (), Tumor_5 had significantly higher than average co-expression with p<0.01. The R package ‘lme4’ (version 1.1.23) was used for the mixed-effects model.

### Phenotype association with mutation signatures

We obtained the molecular variant/mutations of a limited gene panel (Cancer genetics Inc.) in 22 subjects in this cohort and focused the mutation model on influential genes previously reported in lymphoma (20,21). We used the ordered list of somatic copy number alterations (SCNA) reported by Chapuy et.al which identified the five coordinate DLBCL genetic signature clusters C1 (***BCL6, BCL10, TNFAIP3***, *UBE2A*, ***CD70, B2M, NOTCH2, TMEM30A, FAS***, *TP63, ZEB2, HLAB, SPEN, PDL1*) of which our targeted panel overlapped with 8 top ranked mutations in the C1 signature. Similarly, C2 (***TP53***), C3 (***BCL2, CREBBP, EZH2, KMT2D, TNFRSF14***, *HVCN1*, ***IRF8, GNA13, MEF2B, PTEN***), of which the targeted panel overlapped with 9 of the 10 C3 signatures. C2 contained only 1 SCNA of which our panel contained. Our panel overlapped with C4 (***SGK1, HIST1H1E***, *NFKBIE*, ***BRAF***, *CD83*, ***NFKBIA, CD58, HIST1H2BC, STAT3, HIST1H1C***, *ZFP36L1*, ***KLHL6***, *HIST1H1D*, ***HIST1H1B, ETS1***, *TOX, HIST1H2AM, HIST1H2BK*, ***RHOA, ACTB***, *LTB*, ***SF3B1, CARD11***, *HIST1H2AC*), 15 of the top ranked C4 genetic signature. The C5 (***CD79B, MYD88***, *ETV6*, ***PIM1, TBLXR1***, *GRHPR, ZC3H12A, HLAA*, ***PRDM1, BTG1***) genetic signature contained 6 top ranking genetic mutations.

The Chapuy coordinate genetic signature model used the relative TME proportions per core in a multivariate generalized estimating equation model and used the Bonferroni adjustment method by multiplying the p-value by 7 because we used 7 forward selection steps used for identifying significant subsets of CD4 (hypothesis test 1), CD8 (2), macrophage (3), TREG (4), CD4/CD8(5), CD4/CD8/Macrophage (6), CD4/CD8/macrophage/TREG (7) hypothesis tests. The R package ‘geepack’ (version 1.3.1) was used with the exchangeable correlation structure. The Quasi-Information-Criteria was compared with other correlation structures, and they were similar or worse to the exchangeable structure.

Subjects with a variant on a specific gene such as *BCL2, BCL6, CD79b, TP53, SGK1 EZH2, NOTCH1/2*, and *MYD88* amino acid L265P were identified. Co-occurrence between mutations were studied using a Boolean set formation separating mutations that co-occurred or occurred independently. *TP53* (n=8) occurred independently in 3 subjects, and co-occurred with *CD79b* in 4 separate subjects, and once with *SGK1*. Of the 4 *TP53*+*CD79b*+, one subject had an additional variant in *MYD88*^*L265P*^. BCL2 (n=3) co-occurred once as an independent mutation, once with *BCL6* and *EZH2*, and once with only *EZH2. CD79b* variant (n=6) occurred once with *BCL6, 3* co-occurred with *TP53*, 1 with *TP53* and *MYD88*^*L265P*^ and 1 as an independent mutation. *CD79b* did not co-occur with *EZH2*.

> To compute the log-odds (Figure 2B), the estimates were derived using ‘epitools’ R package, to create a 2×2 contingency table of with the rows corresponding to a specific mutation present (+) or not present (−), and the columns corresponding to the counts of cells corresponding to NGBC or GCB. from only the 22 subjects that had mutation screening.

### Estimation of driver proteins on sub-cluster

We avoided calling a sub-cluster immune subset “positive” or “negative” by arbitrary thresholding but used a linear mixed-effects model to derive effect estimates of markers across all sub-clusters. We constructed the linear model using the ROI standardized single-cell expression by mapping all cells per ROI to a standard normal distribution. Hence all ROIs had standardized marker intensity, and for generalizability all estimates excluded non-treated subjects or subjects lost-to-follow-up. We provided point estimates using complete data only. The point estimates and 95% confidence intervals on key markers were derived using (‘lme4 (v.1.1.21)’, and ‘multcomp (v.1.4.12)’ R packages). The point estimates for “driver”, or protein enrichments, on sub-clusters were performed using mixed-effects linear model which treated the case as a random effect, and the sub-cluster mean expression per ROI as the features. We created custom contrasts for all sub-cluster average expression and tested for enrichment by comparing to the global mean for that protein in a single model. The significance of a particular sub-cluster is interpreted as significantly higher/lower to the grand average of that protein (p<0.05), which determined “+/-” for that cluster. The confidence intervals were generated using general linear hypothesis test (‘glht’) function from the R package ‘multcomp (v.1.4.12)’ and derived from ROI standardized values which we hope increases reproducibility in this field.

### Sub-Cluster association with chemotherapy response and refractoriness

The Barnes-Hut t-stochastic neighborhood embedding (tSNE) was constructed using (‘Rtsne (v.0.15)’ Package), and UMAP (umap v0.2.2.1) were used. After performing the UMAP using all single-cells, manual annotations of the sub-clusters were performed by visual inspection. The tSNE parameters were initial dimensions: 50, perplexity : 30, theta: 15. In order to compare enriched/depleted immune subsets, the case proportions for each sub-cluster were derived (%), as well as the standard error of the mean 95% confidence interval (N=33) by averaging ROIs at the case level. The p-values for proportion enrichment were computed using simple linear regression (Figure S9). In order to test enrichment of LAG-3 comparing REF to CR, we fit a One-Way-ANOVA using a mixed effects model and tested pairwise contrasts for each immune subset.

The additional feature “Tumor Spatial Regularity” corresponding to the Clark-Evans aggregation index discussed in Figure 5.

For the neighborhood enrichment comparing refractory to response (Figure 3D and 6A), we performed neighborhood analysis on the full cohort and identified significant interactions using 1,000 permutations which generated the null distribution and a significance threshold of 0.025. For Figure 3D, from the reference of tumor we computed the total significant interactions of all immune phenotypes as a total proportion. The response status relative proportion of significant interactions were depicted along with the 95% confidence intervals for Figure 3. The similar tumor neighborhood summary case proportions were computed similarly for 6B.

### Cross-cohort analysis of DLBCL and Hodgkin lymphoma (HL)

To compare the TME in both diseases, we subset both experiments to include only CD4, CD8, Macrophages, and TREGs, and omitted all other phenotypes present in either experiment. The TME expression of integrated data including both DLBCL and HL identified primary phenotypes in the TME in both diseases. The HL data was obtained from our recent publication in Hodgkin’s lymphoma that included 5 lymph nodes, 22 cases of HL (1 lymphocyte rich, 9 mixed cellularity, and 12 nodular sclerosis subtypes) (31). This experiment was performed with a 36 panel list, of which 21 overlapped with the DLBCL experiment (CCR4, CD206, CD20, CD30, CD3, CD45RA, CD45RO, CD4, CD68, CD8, CXCR3, EphrinB2, FOXP3, HLADR, ICOS, Ki67, LAG3, PD-1, PD-L1, TIM-3, and TBET). The meta-clustering (k_1_=45, k_2_=15) process identified 17 HL meta-clusters using Min/Max normalization as recommended by original authors (36). To compare the TME expression in the 75 ROIs from both diseases, we subset both experiments to include only CD4, CD8, macrophages, and TREGs. Prior data integration, we first selected markers that had minimal differences per ROI across by visual inspection of the ROI interquartile ranges across both diseases. Markers such as CD4, CD8, CD68, FOXP3, CD206, DNA, PD-L1, PD-1, TIM-3, CCR4, CXCR3, ICOS, Ki67, and cell area were selected because they indicated similar inter-quartile ranges and most constant means across experiments. To integrate the single-cell data, we first standardized the expression to-standard normal, and then used ‘limma (v.3.40.2)’ to fit an empirical Bayes linear model and regress out the batch effects related to the 2 experiments at the single-cell level (‘sva’ (v.3.32.1) package). To evaluate the batch effects, we used DNA, and the area of the cell objects as a positive control because we should expect that DNA content, and immune cell areas should be very similar across experiments. After data integration, we observed consistent area sizes across experiments and DNA expression. In contrast, LAG-3 failed to integrate and had dissimilar dynamic range. and was excluded. PCA, at both the case level and single cell level, along with uniform manifold mapping (uMAP) at the single cell level were performed to visually inspect the presence of batch across diseases.

After selecting markers and performing integration, we used the k-nearest neighbor batch-effect correction test (‘kBET’ R package (v.0.99.6)) to quantitatively measure the batch effect. This test is a metric to measure the quality of the batch correction and performs a Pearson’s χ^2^ test comparing a randomly sampled subset to measure the association to the levels of batch covariates (40). The Pearson’s χ^2^ assumes that the data are inter-mixed, and the kBET returns a metric of the average rejection rate of the null hypothesis across the iterations (number of iterations= 100) of sampling. Hence a low kBET score, corresponds with a low rejection of the null hypothesis in the χ^2^ test which assumes that the data are well inter-mixed. Therefore low kBET scores corresponds with low batch effects. The kBET scores across all phenotypes yielded 0.2422 score, and CD4 had the lowest kBET score (kBET=0.1484, p-value=0.31), whereas TREGs had the highest batch effect score (kBET=0.37, p=0.08). Note that we tested for batch on each TME component separately and did not adjust the p-values for multiple test corrections. The input for the kBET metric where the 75 unique ROIs across both experiments (rows) and the columns were the Z-transformed selected protein measures previously described.

We examined the explained variance of the selected integrated expression markers by the phenotype levels (CD4, CD8, MAC, TREG), or the disease type (HL or DLBCL). For a well-integrated experiment, the integrated expression should have a low explained variance associated with disease type. We performed a Two-Way ANOVA regressing the protein expression onto categorical features such as phenotypes (4 levels) or disease type (2 levels). For PD-1, we observed that the phenotype categorical variable was a significant explanatory variable (F-statistic=15.38, p-value=0.025), whereas the experimental factor was not (F-statistic=1.32, p-value= 0.33). Similarly, PD-L1 the phenotype categorical variable was a marginal explanatory variable (F-statistic=4.56, p-value=0.12), whereas the experiment feature was not (F-statistic=0.02, p-value=0.88). For TIM-3, the phenotype feature explained more of the variability of TIM-3 (F-statistic=2.73, p-value=0.22) compared to the experimental factor (F-statistic=0.06, p-value=0.82). The ANOVA indicated that the average expression had more association with the variability across phenotypes levels as opposed to experimental /disease type.

In order to compare proportions of the TME across disease types, the case relative proportions(%) were tested using simple linear regression, and the p-values were multiplied by 4 using Bonferroni multiple test corrections, using significance threshold of 0.05 as the type-1 error rate. We compared the integrated expression of 3 (TIM-3, PD-1, PD-L1) proteins at the patient level average and used a Student’s t-test to compare case average mean differences.

The joint TME unsupervised clusters were annotated (Figure S12C) using threshold calls determined by a mixed-effects linear model (‘lme4’ version 1.1.23) which for each phenotype a contrast was developed which compared for significant differences of a given sub-cluster to the primary parent phenotype average and determined significant above average (+), average expression (mid), or significant below average (−) expression. The annotated heatmap of the labels are depicted in Figure 4A. The cluster proportion enrichment (DLBCL/HL) were measured using logistic regression of the disease type regressed onto the cluster proportions using univariate model, and p<0.01 alpha level threshold.

In order to further investigated the joint TME we used unbiased tertile cutting of each normalized expression intensity relative to the same experiment and compared the relative case proportions to the reactive lymph node using simple linear regression (Figure S14). This supplementary figure avoided comparing directly the two different diseases, and measured differences between each disease to reactive lymph node using an unbiased gating strategy which created tertile cutting within the same experiment. Each subset (e.g. TIM-3+PD-L1+ macrophage) phenotype proportion was measured relative to total primary phenotype (e.g. macrophages) in that cohort. The relative proportions were then compared to reactive lymph node.

### Spatial classification and topology of DLBCL

The algorithm was motivated by Yuan et. Al study of infiltrating lymphocytes in breast cancer using H&E whole tumor slides (55). Yuan defined infiltrating lymphocytes by using the centroid (the average of 5 nearest neighbors (NN) Euclidean distance to cancer cell) which identified a lymphocyte that resided inside a convex hull (domain) of 5 neighboring tumor cells. The 5-NN centroid provided additional information compared to the first nearest neighbor because if a T-cell had shorter distance to the centroid, compared to the 1-NN, this implied infiltration because the lymphocyte was in closer inside the tumor domain. We then sought to develop an algorithm which would classify the tumors by their proximity (5-NN centroids) to nearest immune cells and create a linear ordering by distance. Importantly, the tumor ordering in the context to TME proximity represented a *contour immunographic map*, where furthest distance to the nearest immune cell was analogous to low immune infiltration potential (“steep valley”), whereas tumors that were immediately co-localized to immune cells would have increased immune potential (“top-of-hill”).

In order to develop the algorithm, we generated 4 synthetic point patterns using ‘spatstat (v.1.59.0)’ R package (Figure S14). The synthetic image 1 and 2, had three pattern types, which simulated a germinal center (pattern 1), and the T-cell zone (pattern 2), we included a ‘null’ type (pattern 0) which was included in the distance algorithm, but a shape was left unassigned. The synthetic image 3 had four pattern types. Whereas synthetic image 4 had only two pattern types which demonstrated how the algorithm would order a pattern in terms of distance to the interface (pattern B). We further tested the distance classification algorithm on 6 reactive lymph nodes (Figure S15). From the raw ablation images (Figure S15C) we observed that the CD20 follicle, which captured light/dark zones, was subset (Figure S15A) into sub-types corresponding with nearest distance to PD-1+TFHs and other T-cells in the paracortex region.

The algorithm simply computed the average distance to the 5-NN (centroids) from each B-cell toward the other immune phenotype, and then used Phenograph to meta-cluster (k_1_=45, k_2_=15), the distances into classes which were then ordered. The distance centroids for each tumor cell were used with Phenograph algorithm to classify the tumor cells into clusters dependent on their centroid distance to the immune cells, which provided an immune contour. We observed that the centroid distances were linear to 1-NN neighbors.

### Multivariate logistic regression of topology class abundances in DLBCL

The multivariate logistic regression was performed on the topological relative case tumor proportions (%) that included the IPI scores (Figure 5D, S16A). We compared the Akaike Information Criterion (AIC) from the multivariate regression model to the AIC from the randomized model which permuted the topology labels 250 times to generate a null distribution for the AIC for comparison. Note for the tumor topological multivariate regression tested the multivariate model of all the topology case proportions after adjusting for IPI. We reported the log-odds of the coefficient estimates along with the coefficient p-values.

### Spatial organization of tumor topology in DLBCL comparing COO and reactive lymph node

The Clark-Evans aggregation index is used to measure spatial organization of a point pattern and was performed using the ‘spatstat (v.1.59.0)’ R package. We compared the Clark-Evans standardized indices at the ROI level in DLBCL and 6 lymph nodes and compared them using a Student’s t-test (Tukey test for multiple comparisons).We did not include tumor_h nor tumor_e into the spatial organization model, because their abundances were highly associated with GCB (p=0.057) and NGBC (p=0.026). Tumor classes that were 10% prevalent were selected, and rare topography classes (tumor_a (0.93%), tumor_b (6.9%)) were excluded. By visual inspection, classes: “c” (15.2%), “d” (15.4%), “f” (21.1%), “g” (11%), and “i” (14.9%) were co-localized, however statistical significance was achieved after dropping tumor_d from the spatial organization model, however the trends were still observed.

### Neighborhood analysis

The neighborhood analysis depicted in Figure 6A used the algorithm deposited by Bodenmiller group [(https://github.com/BodenmillerGroup'neighbouRhood)], and significance was determined using 1,000 permutations and interaction distance of 15µm. Significant interactions used p<0.025 to determine a significant signed interaction. The neighborhood pairwise interactions were performed for all pairs, and the significant signed interactions were depicted as a heatmap along with the corresponding mean normalized intensity, association with clinical features, and mutations.

### Multivariate clinical associations between sub-clusters, topological clusters using multivariate logistic regression

The Figure 6A included a multivariate generalized estimation equation model for for each tumor topology only, hence we did not need to adjust for forward selections using Bonferroni adjust methodology. For genetic mutational signature association, we fit the tumor topology case proportions to a generalized estimating equation which is a hierarchical model which controlled for the nested nature of the numerous cores belonging to corresponding cases. We also fit a global mixed-effects linear model to identify if any tumor topologies significantly co-expressed cMYC/BCL2/BCL6 but did not identify cMYC as significantly expressed.

From Figure 2A, we applied an empirical Bayesian linear model (‘limma’ version 3.44.3) on the participant case proportions using the Boolean gating strategy of the most common mutation combinations depicted in Figure 2A. Using the common co-mutations as explanatory features, we applied the linear model and used Benjamini-Hochberg adjustment method (q threshold of 0.05) to determine significant associations between the tumor topologies and common co-mutations.

### Spatial communities and TME quantification of penetration

The neighborhood analysis identified all pairwise interactions (p<0.025) and note that not all spatial interactions are symmetric. In order to describe the tumor topology neighborhoods, we summarized the TMEs from the topology classes, and toward the immune phenotypes. The total sum of the signed interactions in the outer/dispersed (topologies: b, e, g, and h), the tumor periphery (topologies: c, I, f) and tumor core (topology: d) were computed. We defined semi-penetrating as TMEs significantly attracted to the periphery zones and penetrating as TMEs significantly attracted to the tumor core. The 95% confidence intervals were computed using the signed interactions, and the Student t-distribution (d.f.=7).

## ACKNOWLEDGEMENTS/FUNDING

This project was supported by funding from the Ming Hsieh Institute at University of Southern California to A.M., J.H. and P.K. Additional support for this project was provided by K08 CA154975-01A1 career development award to A.M. from the National Cancer Institute, and a Paul Allen Distinguished Investigator award (Frontiers Group; C. Steidl). Access to the imaging mass cytometry system was obtained through the Fluidigm Early Access Program and support from the USC Michelson CSI-Cancer program. The content is solely the responsibility of the authors and does not necessarily represent the official views of the National Cancer Institute or the National Institute of Health.

## Supplemental tables/figures

**Table S1.** Antibody panel. Related to STAR Methods.

| N | Isotopes | Categories | Antibodies | Clones |
| --- | --- | --- | --- | --- |
| 1 | 161Gd | B cells | *CD20 | H1 |
| 2 | 170Er | Pan T cells | *CD3 | poly-C-term |
| 3 | 156Gd | T Helper | *CD4 | EPR6855 |
| 4 | 145Nd | Th1 | *T-bet | D6N8B |
| 5 | 162Dy | Cytotoxic T cells | *CD8a | C8/144B |
| 6 | 163Dy | Regulatory T cells | *Foxp3 | 236A/E7 |
| 7 | 155Gd | Naïve T cells | *CD45RA | HI100 |
| 8 | 173Yb | Memory T cells | *CD45RO | UCHL1 |
| 9 | 167Er | Cytotoxic granule | <sup>+</sup>Granzyme B | EPR20129-217 |
| 10 | 169Tm | M2 Macrophage/APC | <sup>+</sup>CD206 | 5C11 |
| 11 | 159Tb | Macrophage/APC | *CD68 | KP1 |
| 12 | 174Yb | MHC-II, Dendritic cells | *HLA-DR | YE2/36 HLK |
| 13 | 148Nd | Immunoregulatory | <sup>+</sup>ICOS | D1K2T |
| 14 | 150Nd | Immunoregulatory | <sup>+</sup>CD274 (PD-L1) | 28-8 |
| 15 | 151Eu | Immunoregulatory | <sup>+</sup>CD134/OX40 | Poly |
| 16 | 153Eu | Immunoregulatory | <sup>+</sup>LAG-3 | D2G40 |
| 17 | 154Sm | Immunoregulatory | <sup>+</sup>TIM-3 | D5D5R |
| 18 | 160Gd | Immunoregulatory | <sup>+</sup>Vista | D1L2G |
| 19 | 172Yb | Immunoregulatory | <sup>+</sup>PD-L2 | 176611 |
| 20 | 175Lu | Immunoregulatory | <sup>+</sup>CD279 (PD-1) | NAT105 |
| 21 | 142Nd | Immunoregulatory, Th1 | <sup>+</sup>CD183 (CXCR3) | G025H7 |
| 22 | 149Sm | Immunoregulatory, Th2 | <sup>+</sup>CD194 (CCR4) | 205410 |
| 23 | 146ND | Oncoprotein | <sup>+</sup>BCL2 | EPR17509 |
| 24 | 147Sm | Oncoprotein | <sup>+</sup>BCL6 | K112-91 |
| 25 | 164Dy | Oncoprotein | <sup>+</sup>C-MYC-P67 | 9E10 |
| 26 | 158Gd | Transcription factor | <sup>+</sup>pStat3 [Y705] | 4/P-Stat3 |
| 27 | 168Er | Proliferation | <sup>+</sup>Ki-67 | B56 |
| 28 | 143Nd | Stroma | *Vimentin | RV202 |
| 29 | 144Nd | Vascular | *CD31 | C31.3 |
| 30 | 152Sm | Vascular | *CD34 | QBEnd/10 |
| 31 | 166Er | Vascular, immunoregulatory | <sup>+</sup>EphrinB2 | EPR10072(B) |
| 32 | 176Yb | Nuclei | Histone 3 | D1H2 |

\* Phenotypic markers <sup>+</sup> Inducible markers

**Figure S1:**
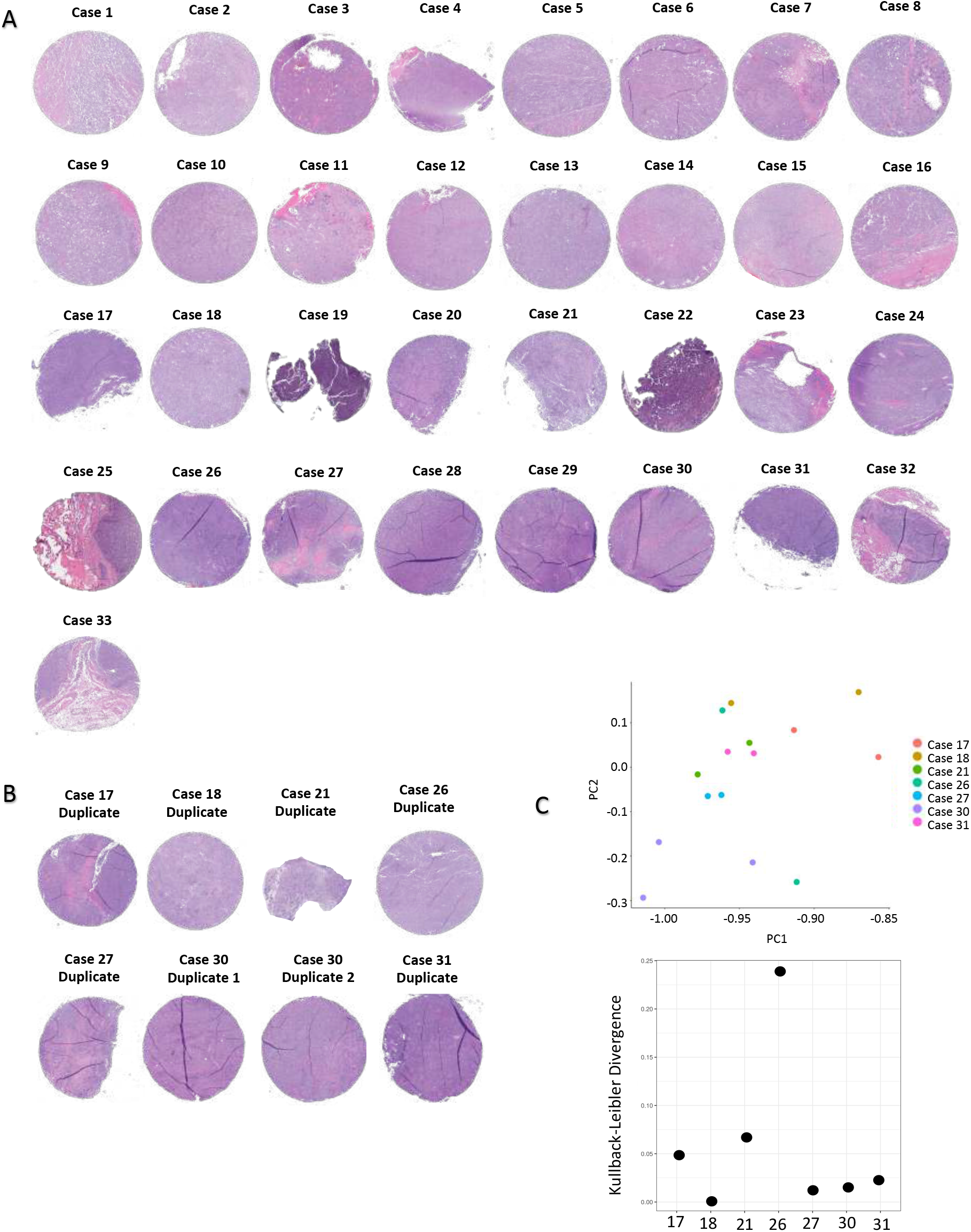
DLBCL cohort. Related to STAR Methods. Images of H&E stained sections of the cases included in the main analyses. A. Images of H&E stained sections of duplicate cores excluded from the main analyses. B. Replicate ROIs H&E. PCA plot shows that the duplicates cluster together, suggesting that small sample areas may be adequate to understand the complexity of tumor immune responses. C. The PCA of the phenotype proportions of replicates (top). The Kullback-Leibler (KL) divergence scores were computed (bottom), and the x-axis denotes the case number. Case 26 replicate ROIs had the highest entropy suggesting that in this case the duplicates are heterogeneous. In this study we averaged the replicates at the case level using a mixed-effects linear model.

**Figure S2:**
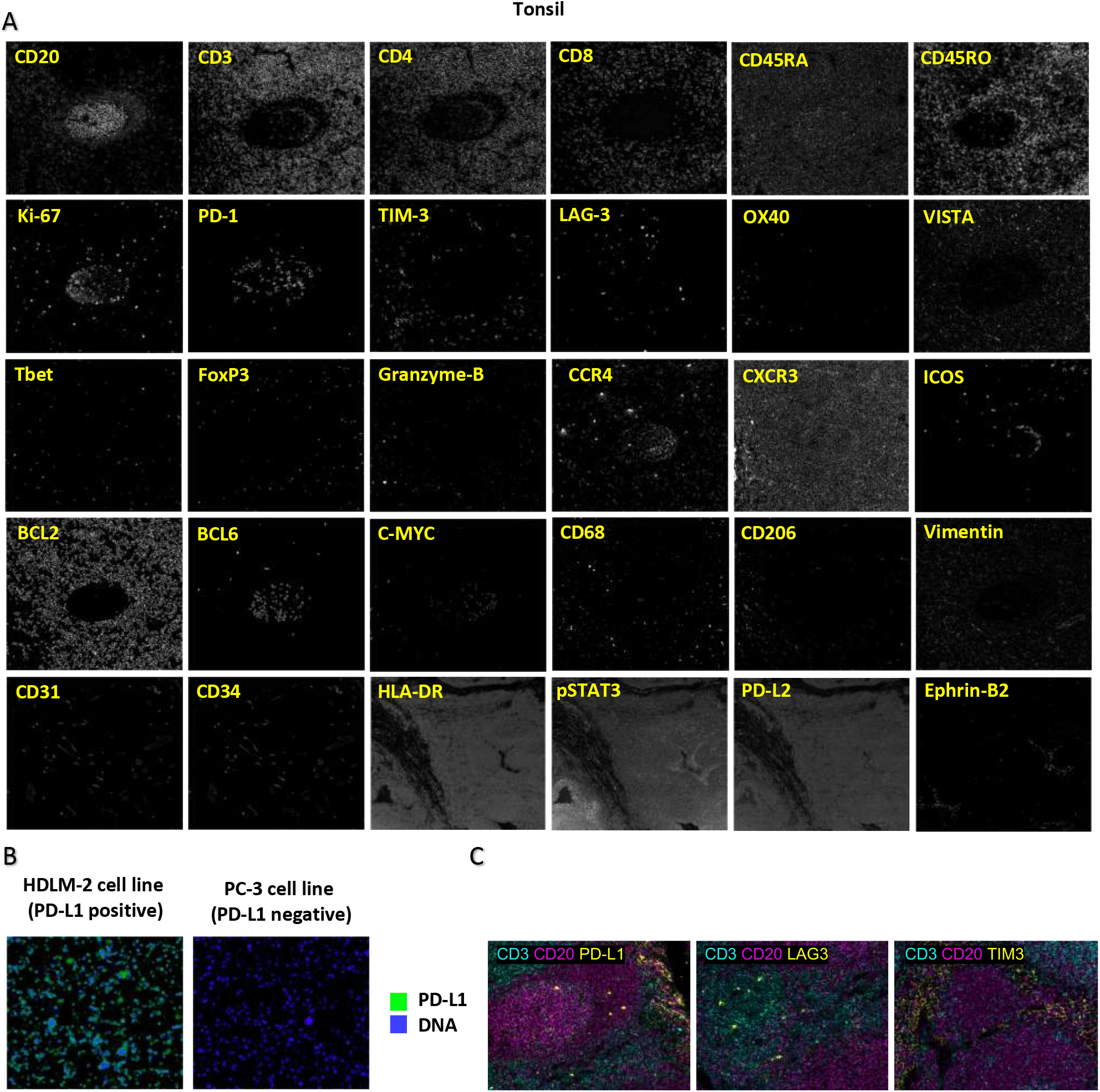
Assay validation. Related to STAR Method. A. A tonsil section was stained with the 32-antibody panel at the same titers used to stain the DLBCL TMAs. B. PD-L1 positivity was validated using HDLM-2 cell line. PC-3 cell line was used as negative control. C. PD-L1, LAG3, and TIM3 antibodies were additionally validated using tonsil tissue.

**Figure S3:**
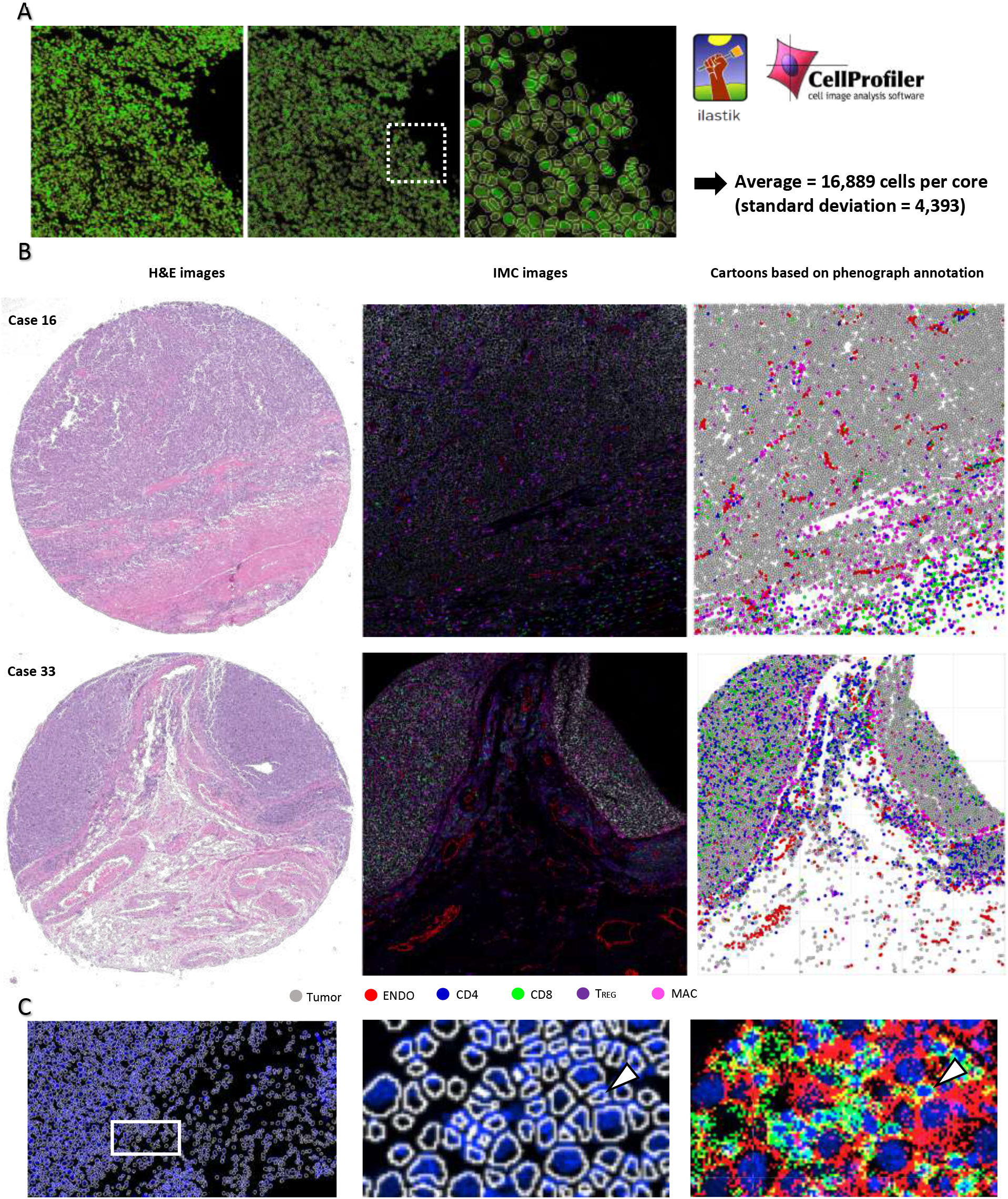
Validation of single cell segmentation. Related to STAR Method. A. LEFT: DNA channel highlighting cell nuclei. MIDDLE: Cell mask overlay. RIGHT: Inset showing perfect mask overlay on each individual cell. Single-cell segmentation yields on average 16,889 cells per TMA core ablated. B. LEFT: H&E images of representative cases 16 and 33. MIDDLE: IMC corresponding images of cases 16 and 33 showing spatial distribution of tumor cells (gray), CD4 T cells (blue), CD8 T cells (green), T_REG_ (purple), macrophages (magenta) and endothelial cells (red). RIGHT: Cartooned images generated based on the segmented masks and Phenograph cluster annotation show matching spatial distribution of the above cell populations. This further highlights the accuracy of segmentation. C. Example of cell in CD4_1 cluster from case 25 (ROI 3967): Arrow = CD4_1 cell, White = segmentation mask, Blue = DNA, Red = CD20+, Green = CD4+, Yellow = CD4+CD20+

**Figure S4:**
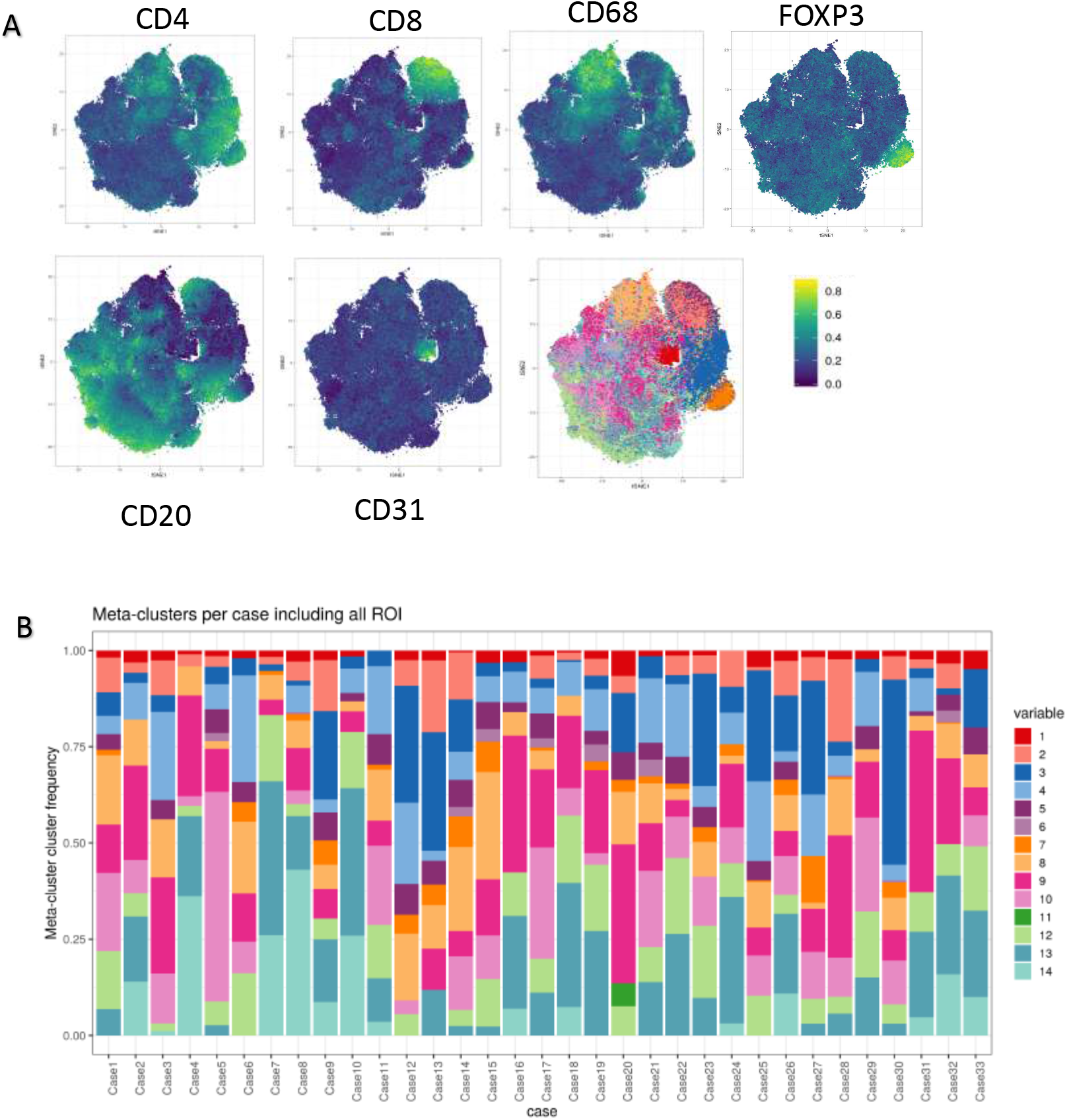
Phenograph clustering and identification of major cell component related to Figure 1. A. t-Stochastic neighborhood embedding (tSNE) graph of the full cohort with major lineage normalized expression maker intensity. The highlighted regions identified major cell components at the cohort level indicates cluster homogeneity for primary lineage markers at the cohort level. B. The case relative proportion of the meta-clusters. We see overall well distributed cluster, without any case specific meta-cluster. The clustering was performed across each ROI, but the centroid (median) expression was used to pool clusters into the meta-cluster level.

**Figure S5:**
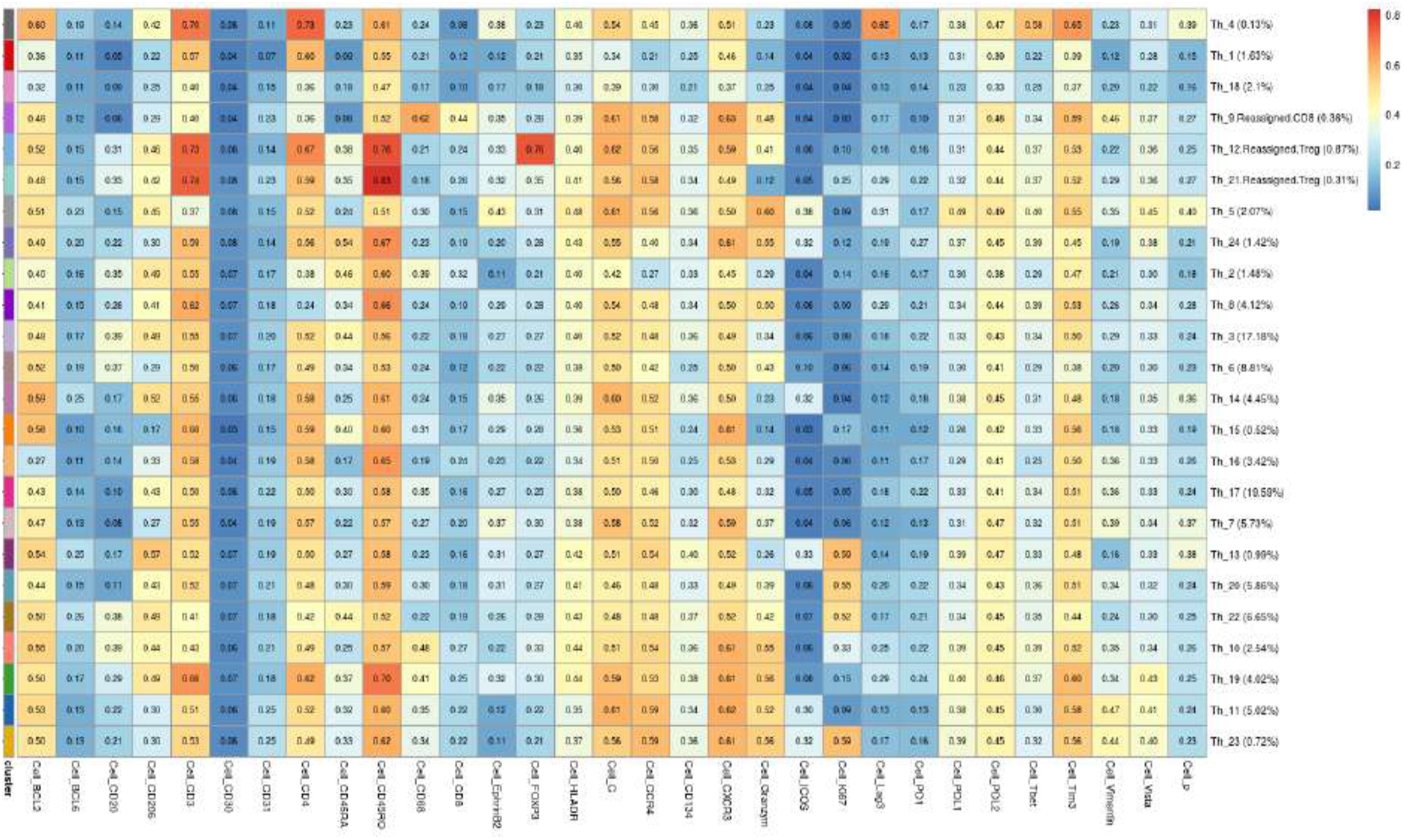
Quality control of CD4 major cell component using sub-clustering re-assignment related to Figure 1. We examined each major cell component (CD4, CD8, T_REG_, B-Cell tumor, endothelial) to ensure that all sub-clusters held homogeneous expression of the lineage expression. We re-assigned any sub-cluster that did not fit the corresponding major component being analyzed. Each major cell component was sub-clustered to optimize homogeneity of lineage specific markers which corresponded with the canonical phenotype of each primary phenotype. CD4 (denoted Th) had primarily uniform expression of CD3 and CD4, with the exception of sub-clusters Th_9 (re-assigned to CD8), Th_12, and Th_21 (re-assigned to T_REG_). Although Th_8 had dim CD4 expression, it did not have over-expression of an alternative lineage marker and was left in the CD4 component.

**Figure S6:**
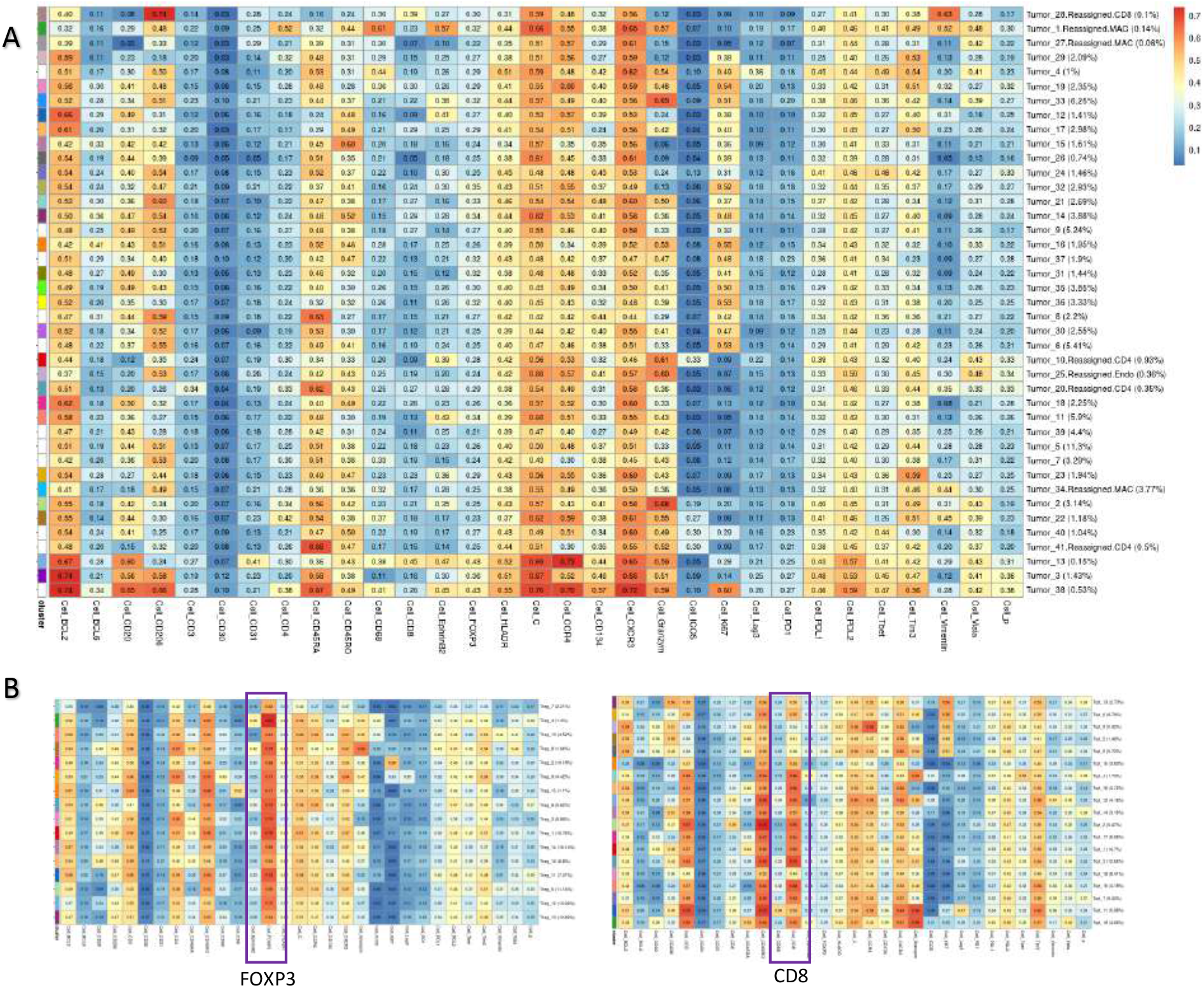
Quality control using sub-clustering re-assignment related to Figure 1. A. We performed quality control analysis on the tumor clusters by sub-clustering each major cell component, and we expected to see uniform expression of the major component marker. (A) depicts the sub-clusters within the “tumor” component, which identified CD20 uniform expression. For the sub-clusters that had dim CD20, they were re-assigned to an appropriate alternative major cell component. The tumor sub-clusters allowed for the re-assignment of ‘tumor’ major cell components with dim CD20, into other major components. B. The T_REG_ (left), and CD8 (right) primary phenotypes did not identify any re-assignments because they had uniform expression of their canonical protein. Similarly macrophages and endothelial components did not require re-assignments.

**Figure S7:**
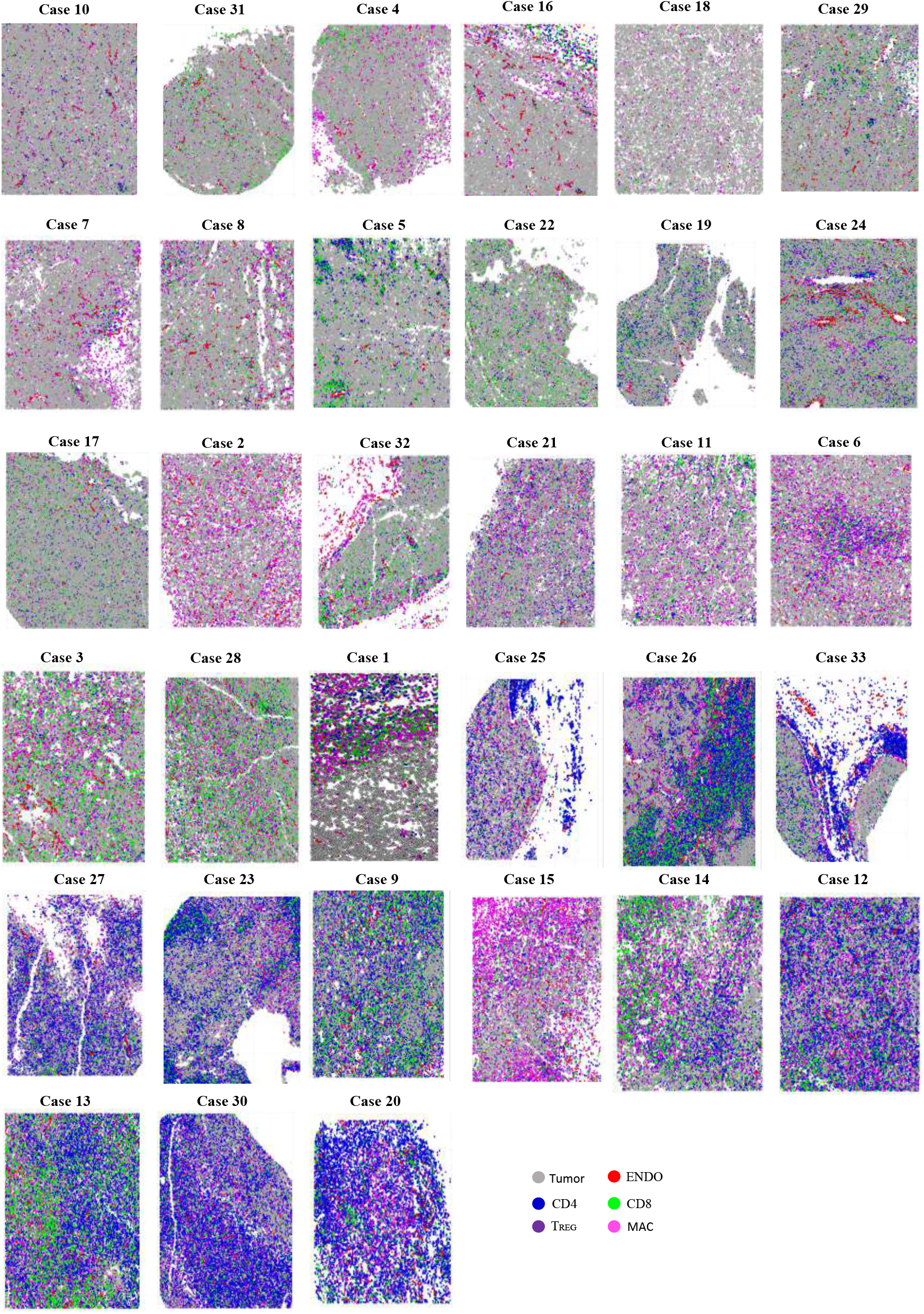
Pseudo-colored cartoons of immune infiltration across cases. Related to Figure 1C. The cartooned images are ranked by increasing immune absolute proportion from 9.42% (case 10) to 90.14% (case 20).

**Figure S8:**
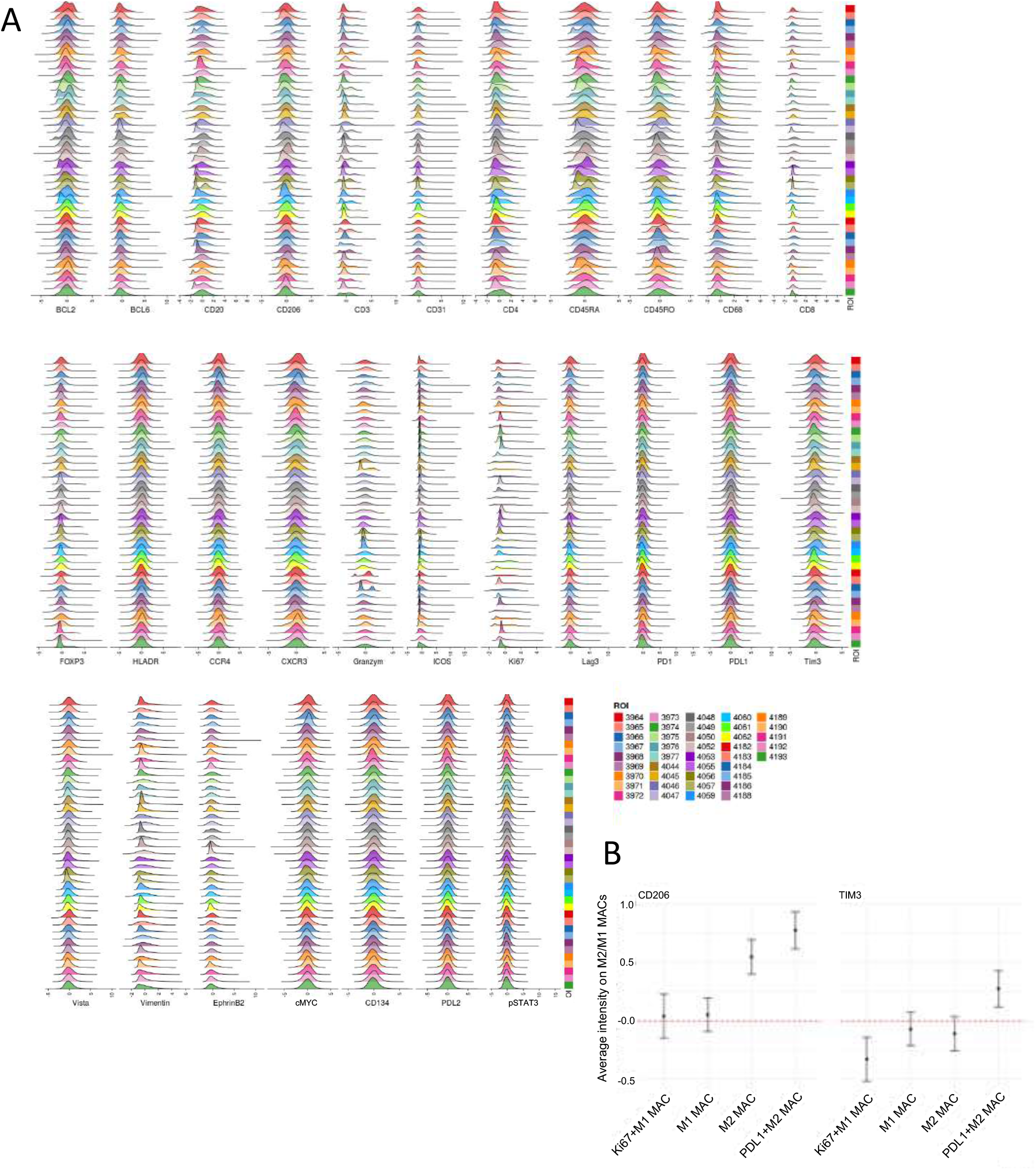
Expression Z-score standardization of each marker across ROI, related to Figure 2. A. For each marker, the min/max normalized scores of all cells were standardized to a Z-distribution across ROIs (I.e. ‘mean normalized intensity’). The x-axis denotes the marker and each y-axis depicts the expression in that ROI. B. The macrophage CD206 and TIM-3 normalized intensity (y-axis) across macrophage phenotype subsets (x-axis).

**Figure S9:**
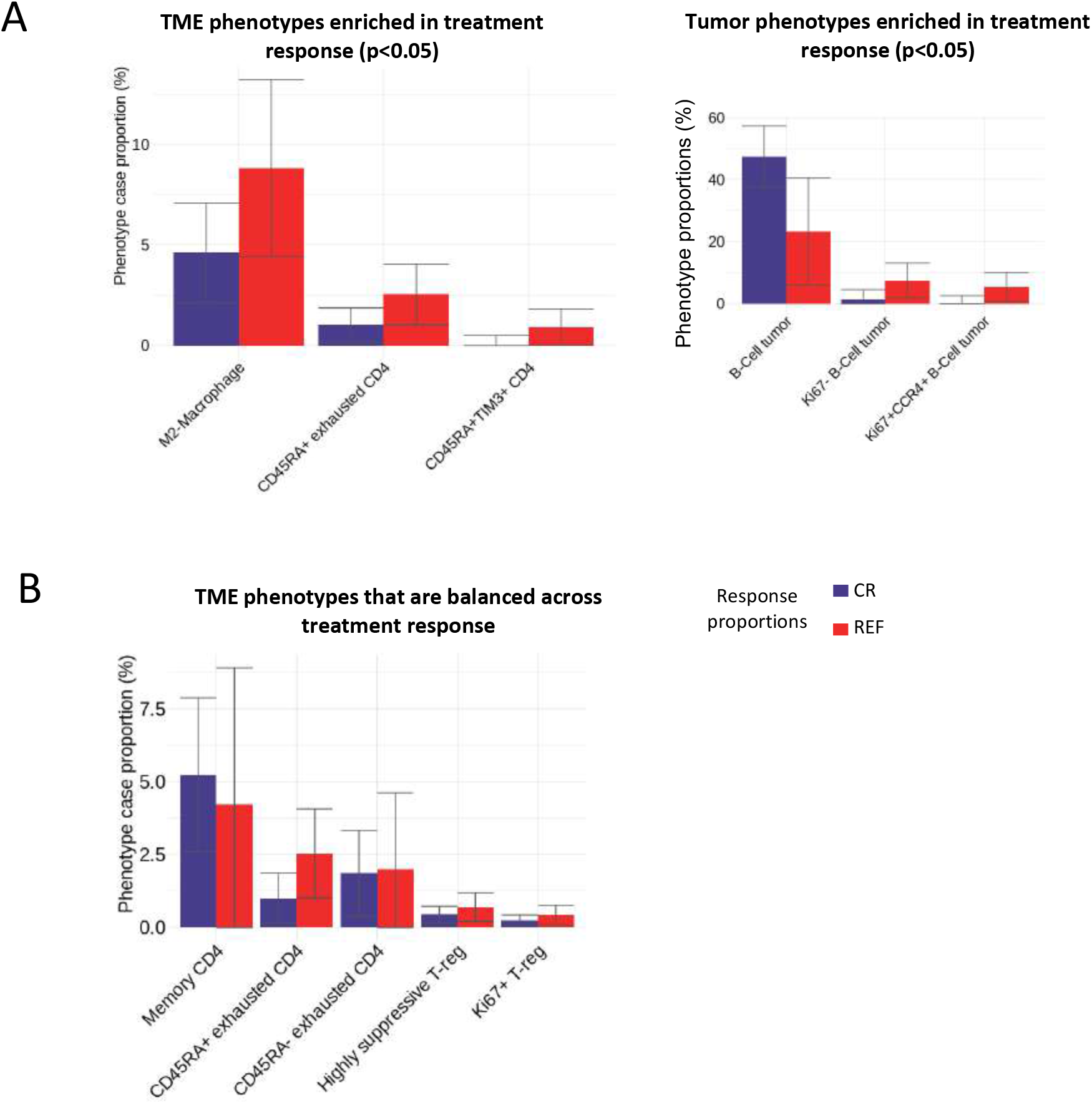
DLBCL tumor phenotype subsets most enriched across treatment response related Figure 3. A. Tumor and immune phenotypes enriched in refractory subjects related to Figure 3. B. The TME phenotypes related to Figure 3C, which show that CD45RA-exhausted CD4 and highly suppressive TREG are proportional across treatment response status but have LAG-3 differentially expressed.

**Figure S10:**
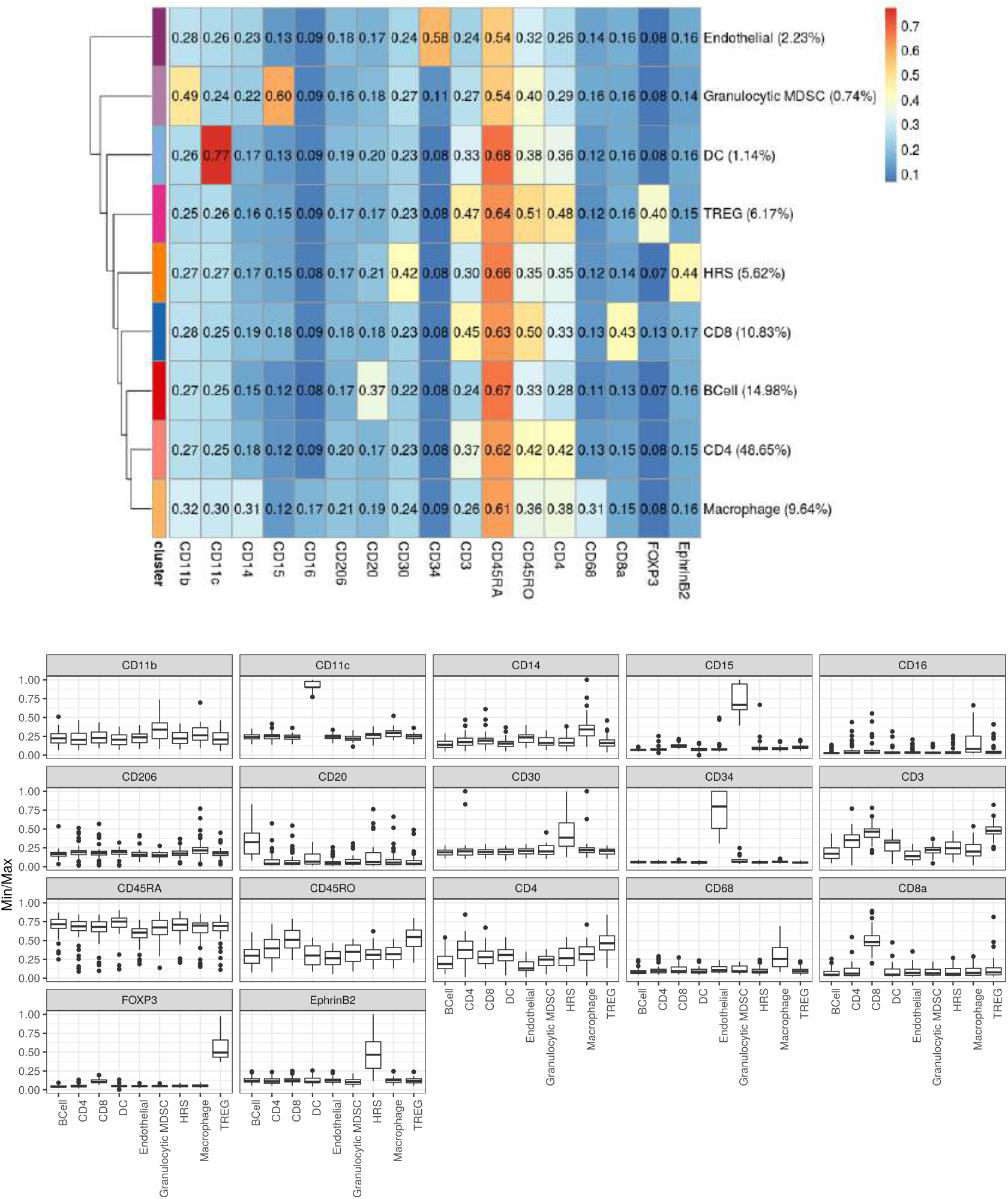
Meta-cluster expression and annotation of major cell components in Hodgkin’s Lymphoma related to Figure 4. The Hodgkin’s lymphoma meta-clustering analysis and annotation of major cell components was performed. Each meta-cluster expression was identified and annotated into major cell lineages. The lineage expression (bottom) corresponds with the canonical phenotype definition; the y-axis shows the Min/Max normalized values, and the x-axis are the phenotypes.

**Figure S11:**
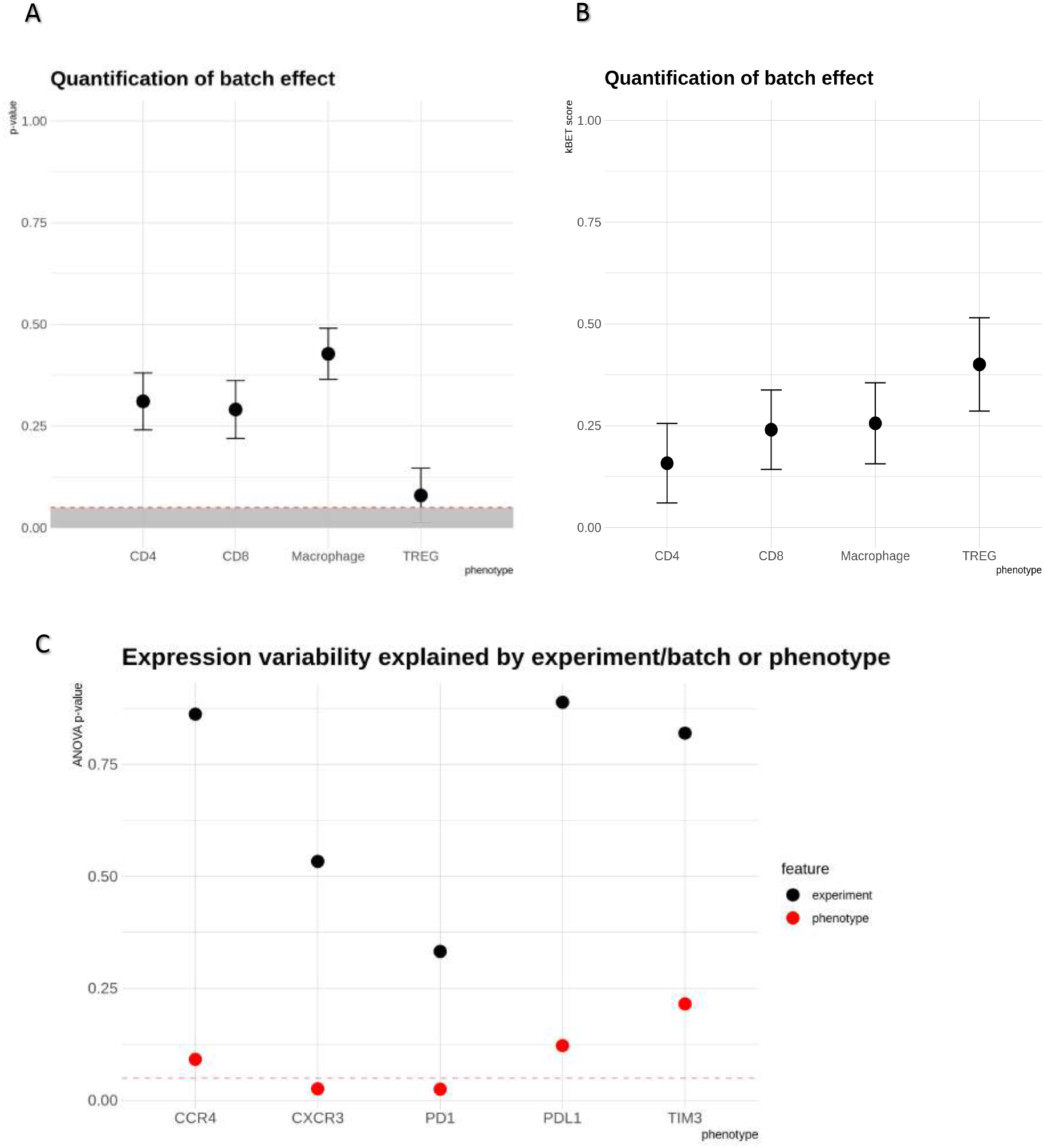
k-nearest neighbor batch correction test (k-BET) scores comparing the TME in DLBCL and Hodgkin’s lymphoma related to Figure 4. A. Using k-BET, the rejection rates and significance level of the Pearson’s Chi-square test are computed. Under the null, the two experiments have no batch effects. The x-axis denotes the TME phenotypes in the 2 experiments, and the y-axis denotes the p-value from the Chi-square test. All the TME phenotypes failed to reject the null hypothesis (CD4 p=0.31, CD8 p=0.49, MAC p=0.43, TREG p-value =0.08). The significance region is depicted in grey. note that we performed the test separately for the 4 phenotypes, but the p-values were not corrected for multiple test corrections. B. The corresponding batch effect scores used in the test corresponding to A. C. The two-way ANOVA regressing marker expression onto the TME phenotypes and the experiment explanatory categorical variables. We can see that the experiment/disease type feature in **black**, (which had 2 levels: HL or DLBCL), had very low mean-square errors (MSE). Whereas the TME categorical variable in **red** (which had 4 levels: CD4, CD8, MAC, TREG) had very high MSE. Hence the expression is more associated with the TME variability and not the disease type.

**Figure S12:**
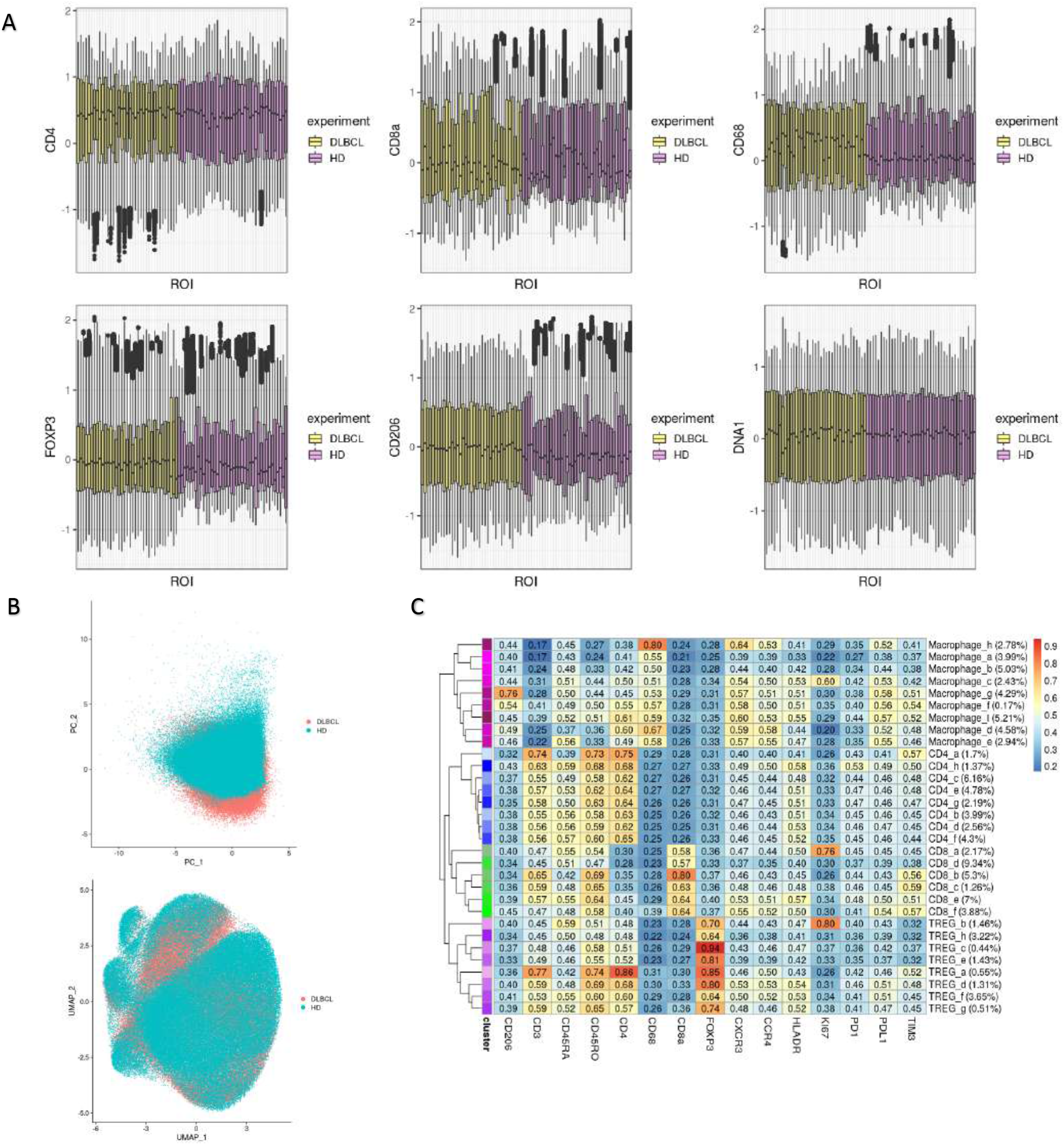
ROI protein expression across the TME in DLBCL and Hodgkin’s lymphoma related to Figure 4. A. Each ROI (x-axis) and the standardized and batch normalized expression (Z-score) across the TME in DLBCL and Hodgkin lymphoma. By visual inspection, we observed similar dynamic ranges with moderate skewness in CD206 and CD68. B. The PCA analysis in Figure 4 was a patient level average of average markers (TIM-3, CD4, CD68, CD8, FOXP3, PD-1, PD-L1, CD206). This PCA and UMAP at the single cell level visually indicate that the batch correction had minimum cohort/experimental bias using the same markers. C. The joint immune phenotype clusters of the DLBCL and HL TME. This was used in supervised annotation of Figure 4A. Positive “+” calls used 0.49-0.5 cut-offs at the cluster level.

**Figure S13:**
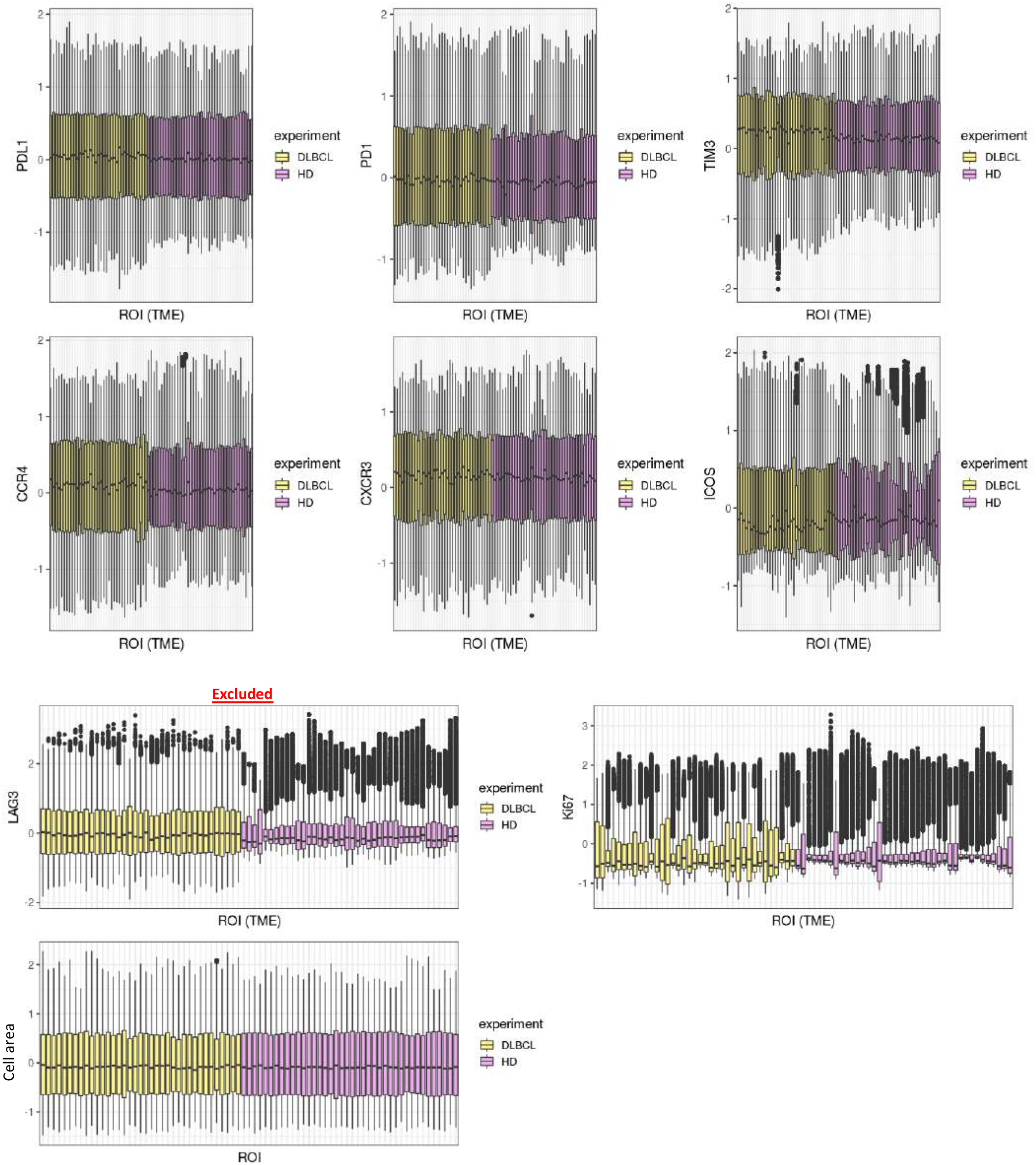
ROI protein expression across the TME in DLBCL and Hodgkin’s lymphoma related to Figure 4. Each ROI (x-axis) and the standardized expression (Z-score) across the TME in DLBCL and Hodgkin’s lymphoma. By visual inspection, we observed dis-similar dynamic ranges of LAG-3, which suggests they had different experimental optimizations, hence LAG-3 was excluded. Ki67 was marginal and was used in the analysis. In both experiments the cell areas of TME were very similar suggesting that the morphological features in both experiments were similar.

**Figure S14:**
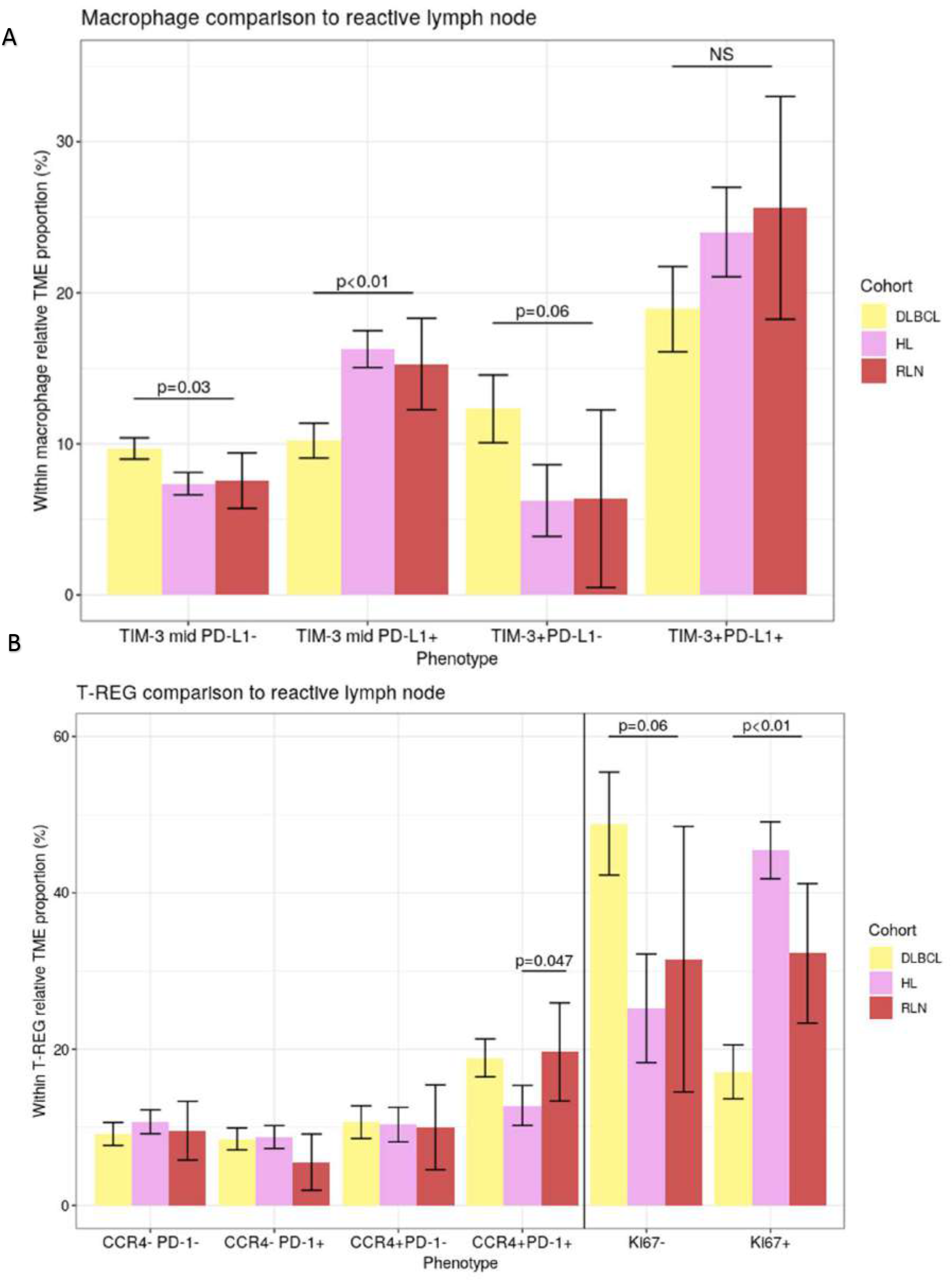
Independent gating strategy of the phenotypes in the HL and DLBCL TME. A. Figure 4A used a linear-mixed effects model to identify significant expression heterogeneity to define phenotype thresholds. We applied tertile gating strategy, which is a common alternative for thresholding phenotypes, and gated each cohort expression relative to its own experiment. The x-axis depicts the tertile gated thresholds of TIM-3/PD-L1 in macrophages found in DLBCL, HL and RLN. The y-axis is the gated proportion relative to the total macrophage proportions found in the TME. The p-values were conducted using simple linear regression with the RLN as the reference. B. Similar to (A) in the context of Ki67 and CCR4/PD-1 gating relative to TREG.

**Figure S15:**
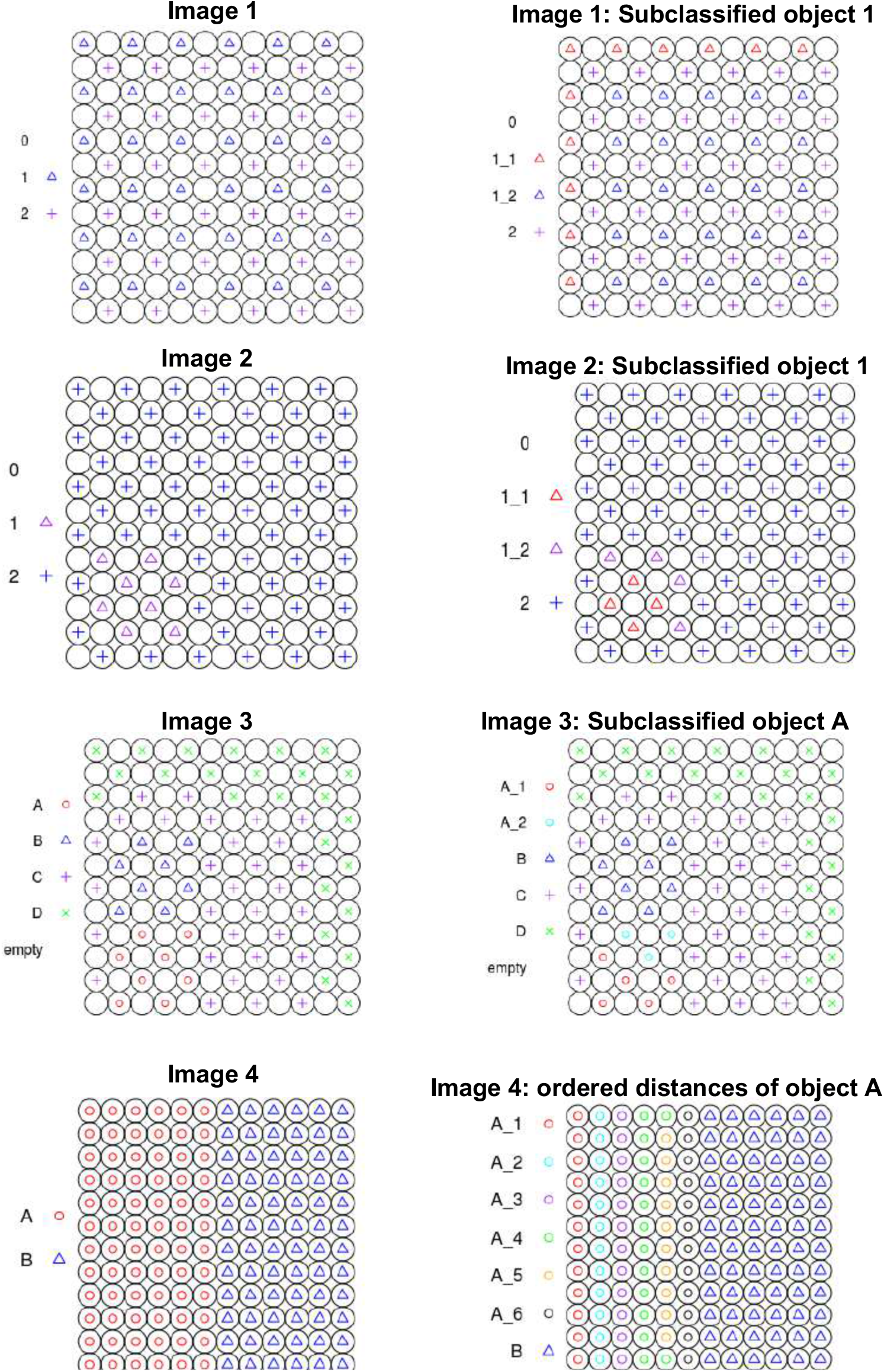
Synthetic cell patterns and spatial sub-classification related to Figure 5. Synthetic spatial arrangement showing how tumor topology model was constructed. We generated 4 synthetic patterns with multiple synthetic cell types. Image1 and Image2 sub-classified object 1 based on distance to other types. Image3 and Image4 identified sub-types for object A based on distance. Image 2 identified the sub-class “1_2” as the “interface” to other object types, whereas class “1_1” was the non-interface class which was determined by their proximity to the other object patterns. Similarly, Image3 identified “A_2” as the interface to other point patterns. Lastly, Image4 identified an ordered distance with the furthest pattern within “A” class to “B” class was “A_1”, and the closest was “A_6”. Image4 demonstrates that the distance classification can order a pattern by distance to the other patterns.

**Figure S16:**
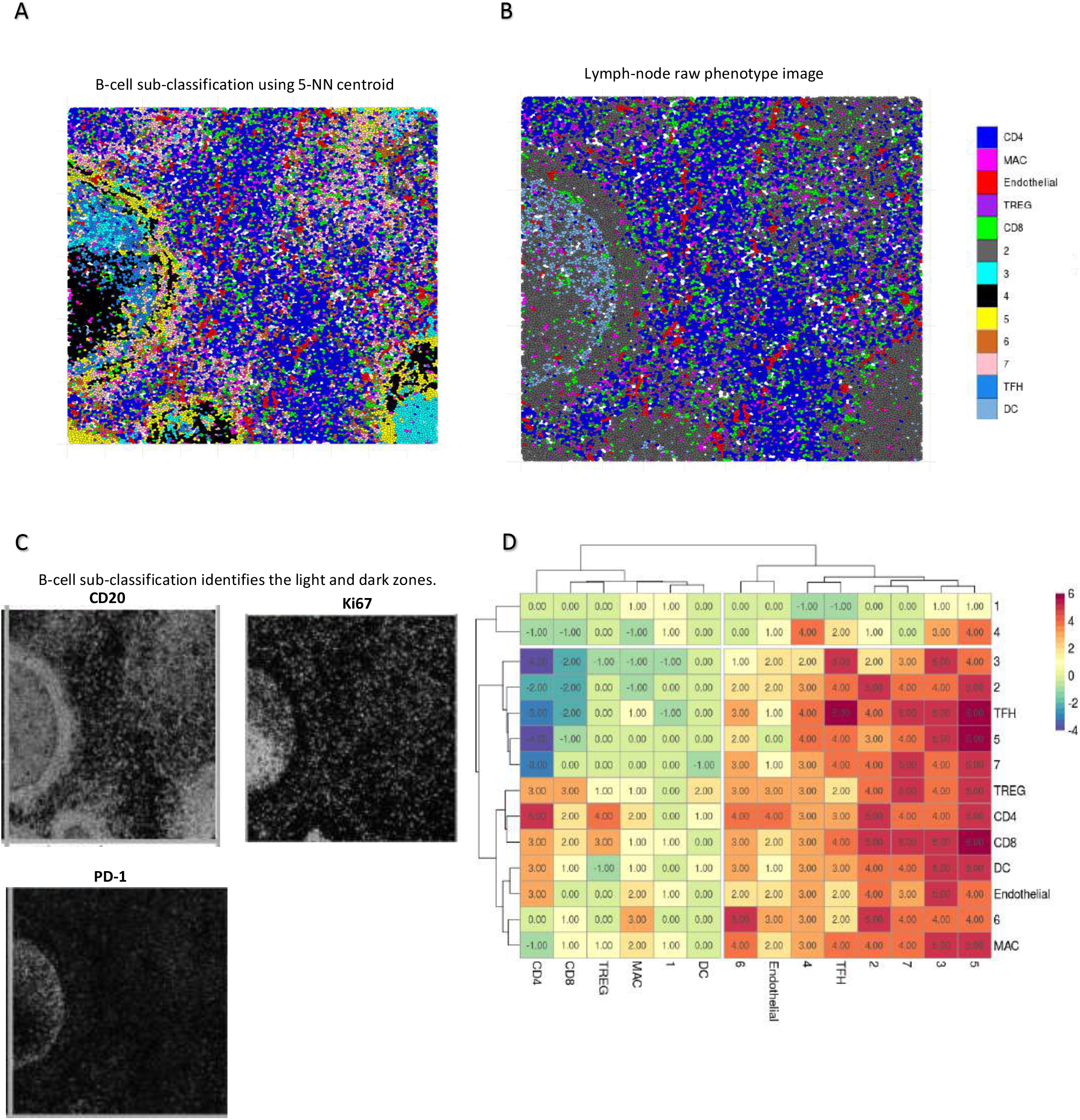
B-cell topology sub-classification related to Figure 5. A. Using 5-NN centroid to sub-classify B-cell major phenotype in the lymph node identified 7 B-cell topography classes based on proximity to other immune cells. Importantly, by clustering in terms of distance to the nearest immune cells (unsupervised), we classified B-cells which identified heterogeneity in the light and dark zones. B. The image rendering of the lymph node showing only the major cell phenotypes annotated. C. The raw ablated images for CD20, Ki67, and PD-1. B-cell sub-classification using the 5-NN centroid was able to identify the heterogeneity of the B-cells in the light (Cluster 3) and dark (Cluster 4) follicular zones. We classified cells based on nearest immune cell. D. The spatial interaction using 1,000 permutations (d=15 microns). We identified that T-follicular helper cells spatially co-localized significantly with B-cell topography classes (4, 3 and 5) from A.

**Figure S17:**
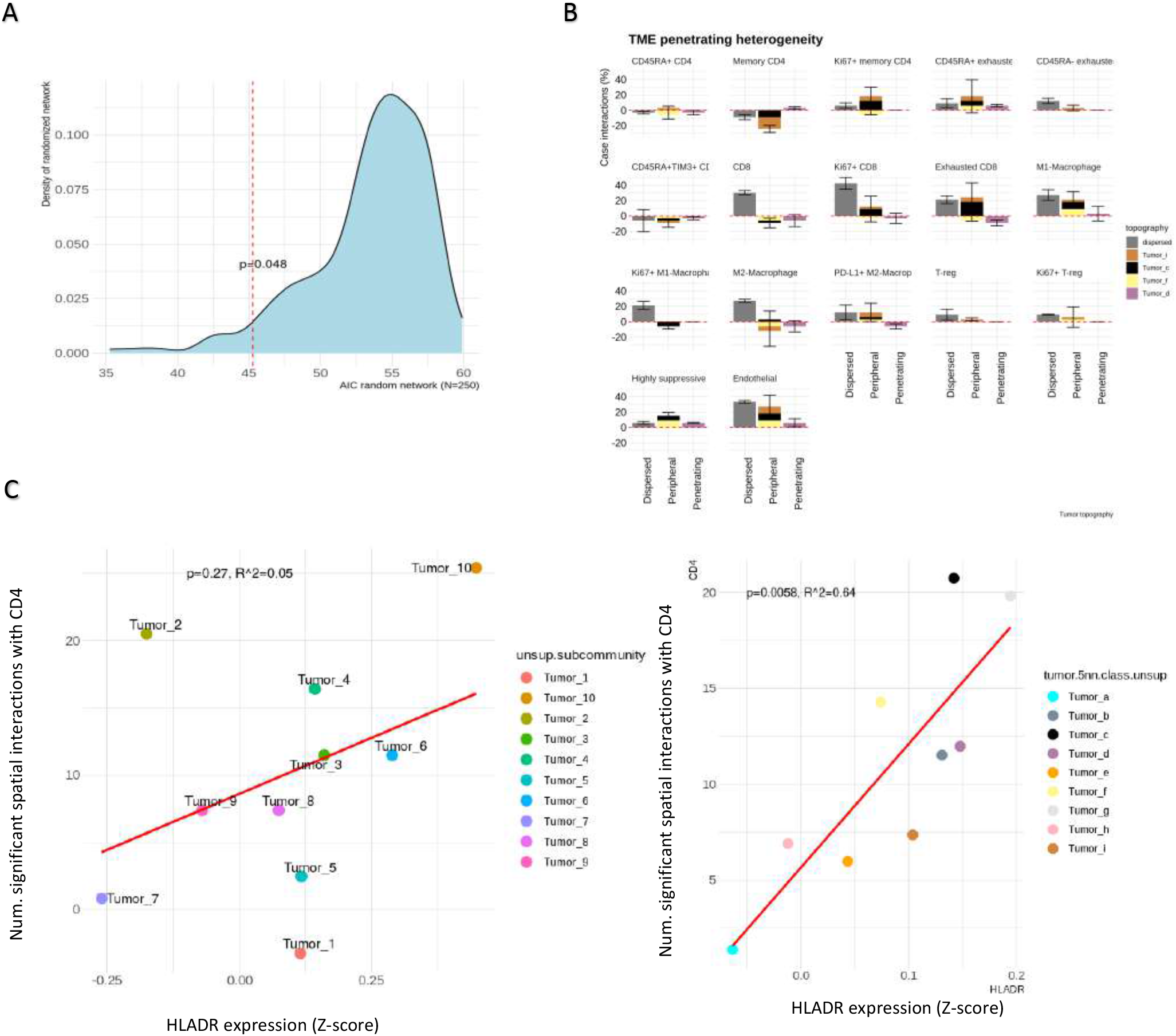
Tumor topography classes analysis details related to Figure 5. A. The logistic regression in figure 5 was compared against a random null model which permuted the topography labels 250 times, and re-performed the logistic regression stratified on the tumor topological cluster labels. The AIC scores were compared between the null model and the observed AIC model to determine robustness. B. Each ROI used 1,000 permutations to determine the spatial interaction (interaction distance=15 microns) from the TME to tumor spatial topology classes. The (y-axis) depicts the total cases with significant interactions (p=0.025) divided by total cases averaged across the tumor topology classes (x-axis). The x-axis depicts the TME based on interactions with dispersed tumors (a, b, e, g, h), semi-penetrating TME by interactions with periphery tumors (c, f, I), and penetrating by interactions with tumor core (d). C. Previously the tumor sub-clusters (Figure 1) were determined by the protein expression alone. Each tumor phenotypic sub-cluster HLA-DR expression (Z-score) is depicted on the x-axis. The spatial interactions (1,000 permutations, interaction distance =15 microns) from the CD4 phenotypes to tumor phenotypic sub-clusters was computed (y-axis). We failed to see a linear correspondence between the tumor sub-clusters HLA-DR expression and spatial interactions with CD4. This is expected because we do not incorporate immune interactions into clustering. The repeated analysis from C was performed, however instead of the tumor phenotypic sub-clusters, we measured the spatial interactions from CD4 to the tumor spatial zones. This analysis indicated that by ordering the tumors by distance to the TME, we see a significant linear correspondence between HLA-DR expressed on the tumor and the number of CD4 interactions.

**Figure S18:**
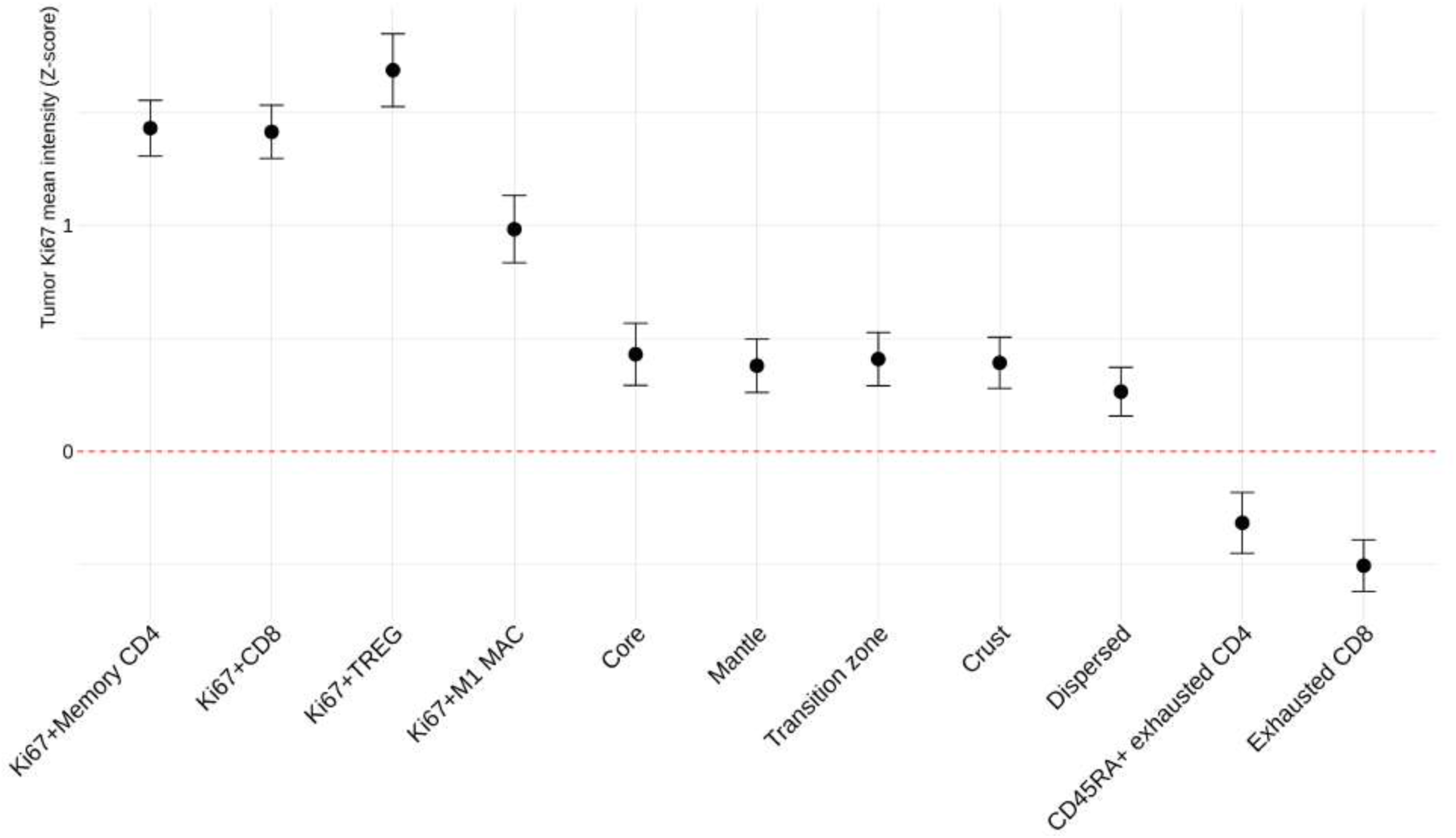
Ki67 expression spatial heterogeneity on B-cell topology related to Figure 6. A global mixed effects linear model across all labeled sub-clusters on Ki67 expression was performed. The depicted plot here shows the estimates of the tumor topology estimated from the global model which identified that the tumor core was moderately proliferative compared to the global Ki67 expression (estimate= 0.43, p<0.001), the mantle (estimate= 0.38, p<0.001), transition zone (estimate =0.41, p<0.001), crust (estimate = 0.39, p<0.001), and dispersed tumors (estimate = 0.26, p<0.001). we observed a decreasing intensity of Ki67 as the tumor zones in the context of TME distance classification. The tumor topologies were mildly proliferative overall, and the tumor topologies were mostly different compared to the dispersed tumor domain.

